# Eligibility for shingles vaccination and hospital-coded dementia in England and Wales: a regression discontinuity analysis in England

**DOI:** 10.64898/2026.07.20.26358345

**Authors:** Fergus Hamilton, Angela Pinot de Moira, Matt Bracher-Smith, Felix Michalik, Siddharthan Chandran, Matias D. Cattaneo, Leandro De Magalhaes, Fernando P Hartwig, David T Arnold, Paul Elliott, Pascal Geldsetzer, Valentina Escott-Price, Bethan Davies, George Davey Smith

## Abstract

We used the September 2013 age-based rollout of the live-attenuated shingles vaccine in England as a natural experiment to estimate the effect of vaccine eligibility on shingles and dementia diagnoses in linked hospital records. Individuals born just before and after the eligibility cutoff were compared using regression discontinuity methods, with follow-up for up to eight years after programme introduction. Eligibility was associated with a clear reduction in hospital-coded shingles diagnoses (RD estimate - 0.12 percentage points, 95% CI -0.153 to -0.079; p = 5.9 × 10^-10^), but there was no evidence of a corresponding reduction in hospital-coded dementia diagnoses (RD estimate -0.06 percentage points, 95% CI -0.40 to 0.27; p = 0.72). Results were robust across denominator definitions, diagnostic-code specifications, estimator choice, placebo cutoffs, and negative-control analyses. The dementia estimate was also close to null in an independently conducted analysis using a separately held HES extract. Comparator analyses in Welsh data with linked primary care and death data did not suggest these results were driven by our reliance on hospital data. These findings do not support a detectable intention-to-treat effect of live-attenuated shingles vaccine eligibility on hospital-coded dementia in England.

## Main

Dementia affects approximately 57 million people globally, with numbers projected to nearly triple by 2050,^1^ yet disease-modifying therapies remain scarce and have modest effect sizes in those who respond.^2,3^ There is therefore substantial interest in preventive strategies, and in particular in whether existing licensed interventions, including vaccines, might reduce dementia incidence through mechanisms beyond their primary indication. Among these candidates, vaccination against herpes zoster (shingles) has attracted particular attention, motivated both by the hypothesis that reactivation of neurotropic herpesviruses contributes to neurodegeneration, and by the broader possibility that vaccines induce trained innate immunity or other non-specific immunomodulatory effects.^4^

The most compelling epidemiological evidence comes from a quasi-experimental regression discontinuity design (RDD) using electronic health records from Wales (the Secure Anonymised Information Linkage, SAIL, databank).^5,6^ This analysis exploited the age-stratified introduction of the live attenuated shingles vaccine (Zostavax) in September 2013, under which people over 80 were always ineligible, while those under 80 received the vaccine when they were eligible. The resulting discontinuity in vaccine receipt at the 1 September 1933 birth date threshold was used to estimate the causal effect of shingles vaccination on subsequent dementia, producing a large but imprecisely estimated 3.5 percentage-point absolute effect (approximately 20% relative risk reduction) with vaccine receipt, largely restricted to women. Similar quasi-experimental analyses in Canada^7^ and Australia^8^ have reported causal effects of broadly similar magnitude, although the sex difference has not been reliably reproduced. Both studies used new dementia diagnoses recorded in primary-care electronic health records as their outcome. Additional quasi-experimental evidence suggested that the vaccine reduces deaths from dementia (in England and Wales^9^); while also having an effect on early stages of dementia such as mild cognitive impairment (only investigated in the Welsh data^10^). A separate analysis in the United States, exploiting the rapid replacement of Zostavax with the recombinant vaccine (Shingrix), reported that Shingrix receipt was associated with further delay in dementia diagnosis (e.g. lengthened lifespan without cognitive impairment) relative to Zostavax.^11^ However Shingrix recipients differed from Zostavax recipients in a wide range of characteristics, requiring the use of propensity score matching in an attempt to control for such differences. The beneficial effect was subsequently suggested to be at least in part related to an adjuvant in the vaccine product, rather than the vaccine itself, although that is contested.^12,13^ In sum, this quasi-experimental evidence suggests shingles vaccination may have an effect across the whole spectrum of dementia; from preventing mild cognitive impairment to preventing death in people with dementia.

Although these findings have been reported across multiple healthcare systems, the underlying sample sizes are modest, and the estimates are therefore imprecise. The magnitude of effect identified in Wales is also at the upper end of what has been seen for any candidate intervention for dementia, and the biological mechanism remains unresolved. Independent replication in a substantially larger dataset, using the same identifying assumption, is required for interrogating the reliability of the reported effects, together more precise effect estimation if such prove to exist.

We performed an RDD analysis in Hospital Episode Statistics (HES), an administrative dataset covering all National Health Service (NHS) inpatient and outpatient activity in England.^14^ Because the date of birth eligibility rule that defined the live shingles vaccination rollout in Wales also applied to the rest of the UK, and because the SAIL RDD assumptions are expected to hold equally in England, HES provides a natural setting for replication. Our primary analyses used HES hospital-coded outcome counts with HES hospital-activity denominators for 6,340,214 individuals born within eight years of the September 1933 threshold, approximately 18 times the size of the original SAIL cohort.^5^ We also used Office for National Statistics (ONS) population denominators as secondary denominator checks. We estimated the intention-to-treat effect of vaccine eligibility on hospital-coded shingles and dementia in England, and then accessed SAIL to test, both theoretically and empirically, whether specific features of either dataset could plausibly explain any difference in results.

We find a clear and robust expected protective effect of eligibility on hospital-coded shingles in line with randomized controlled trial data, demonstrating the reliability of our identifying procedure. However, we did not identify a robust effect on hospital-coded dementia. Analyses in SAIL indicate that none of the candidate biases in our dataset, namely incomplete exclusion of prevalent disease, a hospital-contact sampling frame, or reliance on secondary care coding, are of sufficient magnitude to explain the discrepancy between SAIL and HES.

## Results

Our primary dataset was derived from HES using hospital activity across the study window needed to define the pre-index baseline period and the post-index follow-up period around the introduction of the vaccine on 1 September 2013. HES is constructed by assigning each hospital admission or outpatient activity as an event, which is date-stamped, International Classification of Disease version 10 (ICD-10) coded, and assigned to individuals via their NHS Number. This allows tracking of hospital activity for individuals across the study window. For each individual, we have age, sex, and year and month of birth (exact date of birth was not available in our primary HES extract). For our primary analyses, we had access to HES records from 1 April 2012 to 31 March 2022, a ten-year period largely occurring after the introduction of the vaccine. In parallel, the SAHSU team independently analysed a separately held, aggregated HES extract with a longer pre-programme lookback and week-of-birth resolution.

The vaccine was introduced on 1 September 2013; and therefore we observe, for the whole of England, all NHS hospital events for the 17 months prior to this date, and the ∼8.6 years after. For our RDD analysis, we limited our analytic sample to participants born within 96 months (8 years) either side of September 1933 (September 1925 to September 1941). Because our HES extract records year and month of birth but not exact day of birth, the running variable was defined in monthly birth bins around September 1933. We included individuals born during September 1933 as the vast majority of these would have been eligible. Individuals born after September 1933 were eligible for the 2013 catch-up shingles vaccination programme, whereas those born before September 1933 were not.

### Differences in HES and ONS reference populations

The analytic population therefore ranged from people aged 71 on 1 September 2013, to those aged 88. This population included 6,340,214 individuals. 45.1% were men and 54.9% were women, with a mean age of 78.8 years at the September 2013 programme threshold, as the population is slightly larger at younger ages. To enter the analytic population, participants had to have hospital activity somewhere within the broader extract provided by NHS England in this HES extract (e.g. 2012-2022). Previous studies have suggested that hospital exposure in this age group is extremely common,^15,16^ suggesting that our analytic population will not be dissimilar to the true total population for each age-month bin, but we assessed this by comparing our population estimates to those provided by the Office for National Statistics (ONS). See **Methods** for details of construction of these denominators. The correlation of population size across age month-bins between ONS and HES was extremely high (Pearson’s *r* = 0.996, **Supplementary Figure 1**), tracked across all month bins, and showed no visible discontinuity at the shingles vaccine policy threshold. However, HES estimates were consistently about 32% higher than ONS estimates, likely due to these reflecting transient populations (e.g. travellers from outside the UK), and failure of deduplication (e.g. the same person presenting to different hospitals but being recorded twice), although differences were larger than that observed in other comparable studies.^17^ Nevertheless, after mean-shift correction - calculating the mean percentage difference across age-month bins and deflating HES estimates across all bins by that proportion - the two series were very similar. We therefore used the HES hospital-activity denominator as primary because it is defined in the same administrative system as the outcomes and date-of-birth running variable and report the ONS-denominated analyses as sensitivity analyses. Because both denominator series are smooth and highly correlated across the RD threshold, the choice of denominator will little affect RD identification; it does, however, change the scale on which absolute risks are expressed. **Supplementary Note 1** discusses this choice further.

### Vaccine eligibility strongly reduces hospital-coded shingles diagnoses

We first tested whether we could identify effects of eligibility of the shingles vaccine on hospital-coded shingles. About 90% of clinical cases of shingles are seen in primary care,^18^ so the presence of this code in hospital inpatients reflects both severe cases and incidental cases identified during a hospital admission. Outpatient clinic assigned codes for shingles were extremely rare (6 total across the dataset).

We used a RDD as described in the **Methods**. For the primary analysis, we used rdrobust, an R package for performing RD which automatically selected the mean-squared-error (MSE) optimal bandwidth - that is, the amount of time before and after the discontinuity used to fit the local regression lines - of 21.7 months either side.^19^ We calculated monthly counts of first post-index hospital-coded shingles events using ICD-10 code B02. In our primary analysis we excluded people who had a code of shingles in the prior period (n = 4,575; 0.07% of the HES hospital-activity population). We recorded 27,215 first post-index shingles events over 8 years of follow-up, corresponding to a HES-denominated cumulative risk of about 0.43%. The estimated intention-to-treat effect of vaccine eligibility at the cutoff was -0.12 percentage points (95% CI -0.153 to - 0.079 percentage points; p = 5.9 × 10^-10^, **Figure 1**), interpreted as an absolute reduction in hospital-coded shingles risk. The corresponding secondary ONS-denominated estimate was -0.16 percentage points (95% CI -0.205 to -0.112; p = 2.8 × 10^-11^).

**Figure 1:**
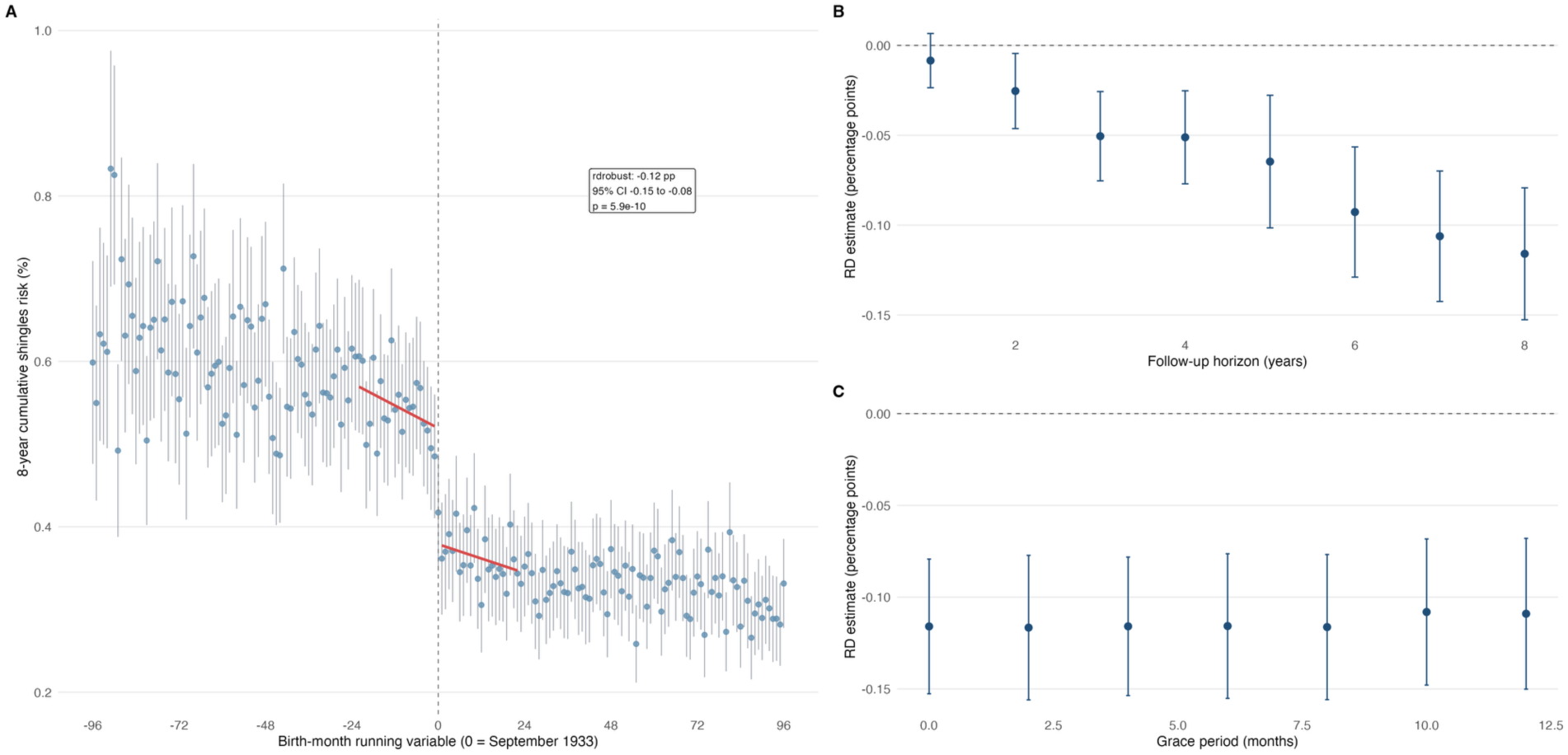
Effect of eligibility for the shingles vaccination on incidence of shingles. Panel A shows monthly mean outcome risks at follow up with 95% binomial confidence intervals and local-linear fits estimated separately on each side of the eligibility threshold, using HES denominators. The outcome is first hospital-coded shingles diagnosis within 8 years after excluding people with a prior hospital-coded shingles diagnosis. Solid lines are weighted local linear fits on either side of the cutoff, restricted to the rdrobust MSE-optimal bandwidth. Panel B shows how the effect estimate varies by increasing follow-up, while Panel C shows the influence of introducing a grace period after the index date before outcome counting begins

Sex-specific analyses showed similar reductions in shingles diagnoses in men and women. We performed several sensitivity analyses, including: (i) restricting outcomes by diagnosis position within HES - e.g. restricting to the primary and secondary diagnostic code; any diagnostic code, or excluding primary or secondary diagnostic codes; (ii) using fixed bandwidths around the RDD period (12 or 24 months) and an alternative regression discontinuity estimator (the R package RDHonest^20^); (iii) testing annual placebo cutoffs at shifts of ±12, ±24, and ±36 months around the true September 1933 threshold - these replicate the RD analysis on a date distant from the actual RD date and are tested as negative control exposures; and (iv) using ONS rather than HES denominators. The shingles discontinuity remained negative in men and women, across diagnosis-position definitions, after excluding the September 1933 boundary month, under the main alternative bandwidth and estimator specifications, and when using the secondary ONS denominator; all placebo cutoffs were compatible with the null (**Supplementary Table 1; Supplementary Figures 2 and 3**).

### Shingles vaccine eligibility did not materially inffuence dementia diagnoses

We then identified dementia using the same codelist as used in the initial RDD analysis.^5^ In the primary analysis, we excluded individuals with evidence of dementia before baseline in HES (152,246, 2.4% of participants excluded); however, first post-index hospital-coded dementia may still include some long-standing but previously unrecorded disease rather than wholly incident diagnoses. This should be less problematic for the regression discontinuity design than for conventional observational analyses, because any residual misclassification would not be expected to change discontinuously at the September 1933 eligibility threshold (see **Supplementary Note 2** for further discussion).

In the primary non-prevalent cohort, 843,772 first hospital-coded dementia events were recorded over 8 years of follow-up, corresponding to a HES-denominated cumulative risk of about 13.6%. We found no robust evidence that eligibility for shingles vaccination influenced hospital-coded dementia (**Figure 2**). The estimated absolute effect was - 0.06 percentage points (95% CI -0.40 to 0.27 percentage points; p = 0.72), corresponding to an effect close to zero. The automatically selected rdrobust bandwidth was about 26.5 months on each side of the cutoff. The corresponding secondary ONS-denominator estimate was similarly null (−0.11 percentage points, 95% CI -0.63 to 0.41; p = 0.68). A narrower alternative dementia definition (from^21^) was also null (−0.02 percentage points, 95% CI -0.38 to 0.33; p = 0.91). Relative to the observed 8-year HES-denominated dementia risk of 13.6%, the primary point estimate is therefore about 0.4% of the observed cumulative risk, with the confidence interval spanning roughly a 3% relative decrease to a 2% relative increase.

**Figure 2:**
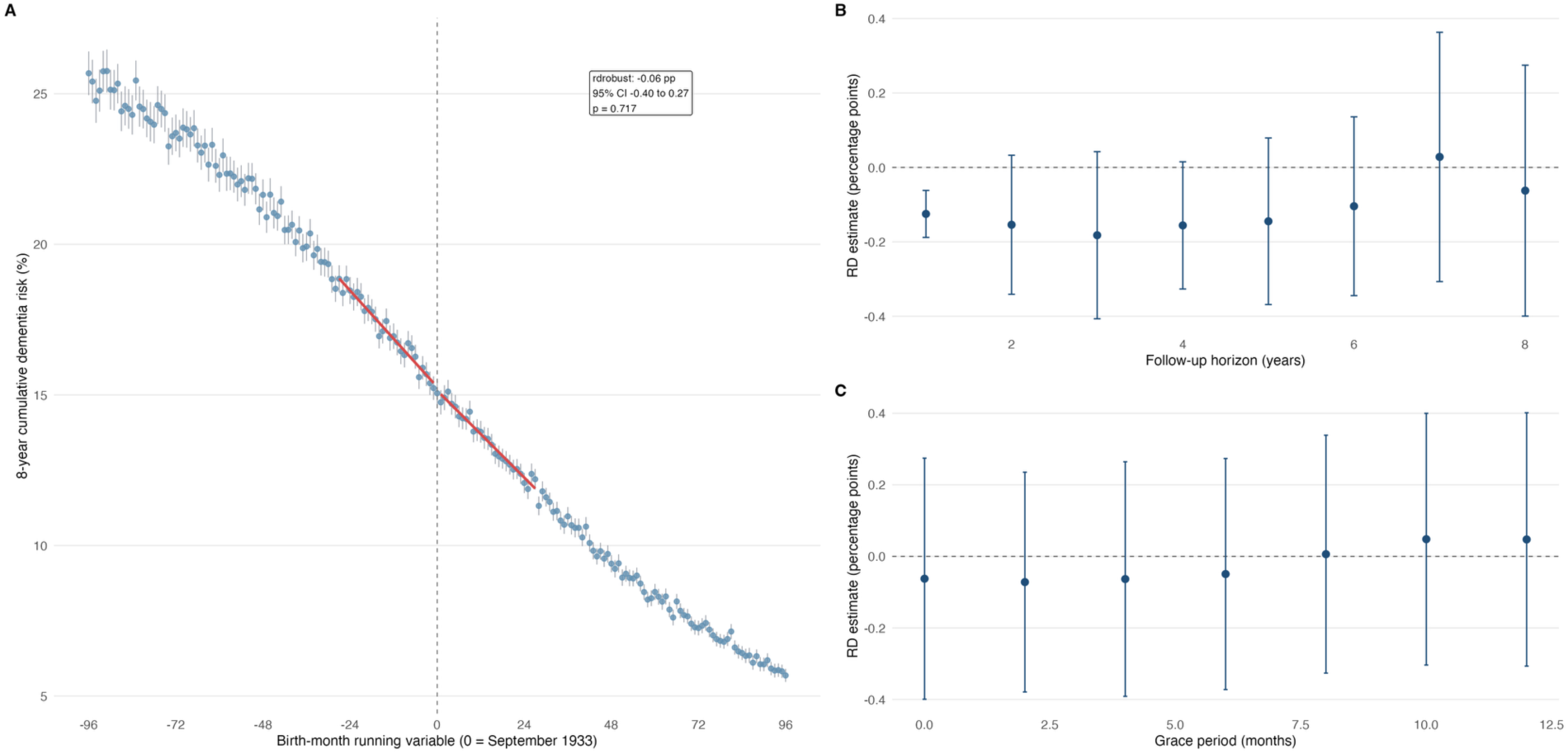
Effect of eligibility for the shingles vaccination on incidence of dementia. Panel A shows monthly mean outcome risks with 95% binomial confidence intervals and local-linear fits estimated separately on each side of the eligibility threshold, using HES denominators. The outcome is first hospital-coded dementia diagnosis within 8 years after excluding individuals with a prior hospital-coded dementia diagnosis. Panel B shows the effect of increasing follow-up, while Panel C shows the effect of introducing a grace period after the index date before outcome counting begins.

Sensitivity analyses were broadly null. Sex-stratified analyses, shown together for shingles and dementia in Figure 3, did not suggest that the dementia null masked a protective effect in either men or women. Results were similar across diagnosis-position definitions, with ONS rather than HES denominators, and under fixed-bandwidth and alternative-estimator analyses. Excluding the September 1933 boundary month also left the result null (−0.03 percentage points, 95% CI -0.37 to 0.31; p = 0.85; **Supplementary Table 2** and **Supplementary Figure 2**). Annual placebo-cutoff analyses at ±12, ±24, and ±36 months were null (**Supplementary Figure 4**).

**Figure 3:**
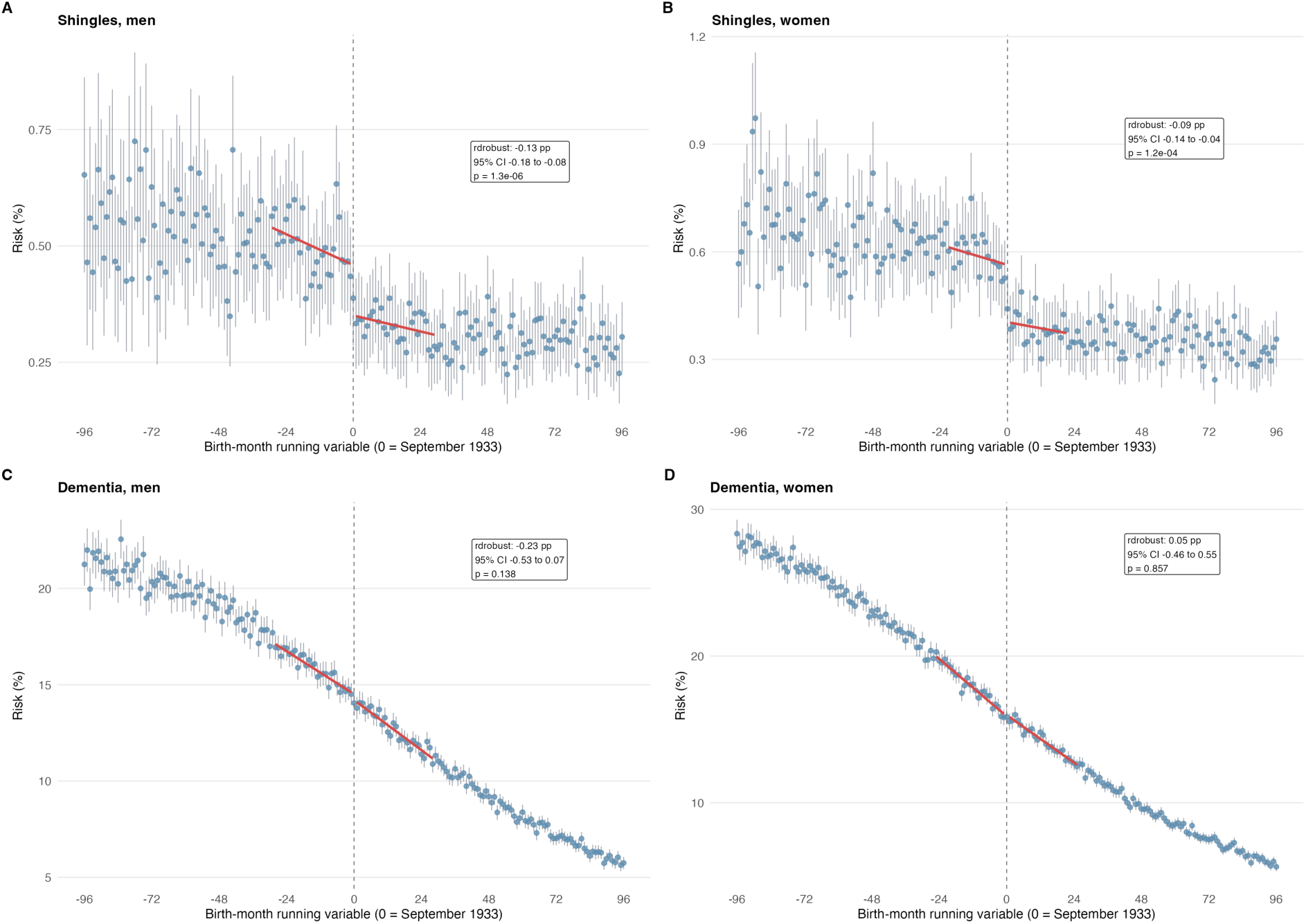
Sex-specific RD analyses for shingles and dementia. Panels A-B show first hospital-coded shingles diagnoses in men and women; panels C-D show first hospital-coded dementia diagnoses in men and women. Points show monthly mean risks with 95% binomial confidence intervals and red lines show local-linear RD fits estimated separately on each side of the eligibility threshold, using HES denominators.

### Early follow up signals are too early to be plausibly caused by vaccination

The main departure from the null pattern was the 1-year follow-up analysis, which showed a nominal protective association (−0.12 percentage points, 95% CI -0.19 to - 0.062; p = 1.1 × 10^-4^), before attenuating at longer follow-up. The confidence intervals around these estimates varied because of the automatically chosen bandwidth in rdrobust.^20^ Importantly, some cases recorded early during the follow-up period will reflect dementia cases that precede the RD cutoff but are observed after it, as the average delay between primary care diagnosis and hospital-coded dementia is approximately 1.6 years.^22^ Therefore, the early signal is likely to reflect prevalent cases first observed in hospital data rather than new incident cases. Additionally, official United Kingdom Healthy Security Agency programme data^23^ show a strong temporal and cohort pattern: in the first programme year, vaccination was offered to the routine 70-year-old and catch-up 79-year-old cohorts. Most vaccinations occurred in the first few months, and coverage in both cohorts exceeded 45% by the end of January 2014. Separately, the interval between recorded vaccination and dementia diagnosis in SAIL showed that dementia had been recorded before vaccination for a proportion of vaccinated individuals (**Supplementary Figure 5**).

We therefore performed a specific analysis with a 12-month grace period (e.g. follow up starting in September 2014) and a fixed 12-month eligible-side bandwidth, while allowing rdrobust to select the older-side bandwidth (e.g. only allowing those aged 79 to be compared to the older, non-vaccinated cohort). This better aligns the analysis with the subgroup plausibly exposed during the first programme year and allows time for vaccination to affect hospital-coded dementia, which would still require an extremely rapid biological effect (∼6 months). In this analysis, we saw no evidence of a dementia signal at any post-grace follow-up horizon (**Figure 4**). We also repeated annual placebo-cutoff analyses for the 1-year endpoint at ±12, ±24, and ±36 months (**Supplementary Figure 6**). The true cutoff retained a nominal early protective association, but a nominal signal was also present at the -12-month placebo cutoff, further suggesting the early protective signal is not specific. As such, we feel this 1-year result is not supportive of an early signal, rather it reflects the play of chance across numerous sensitivity analyses.

**Figure 4:**
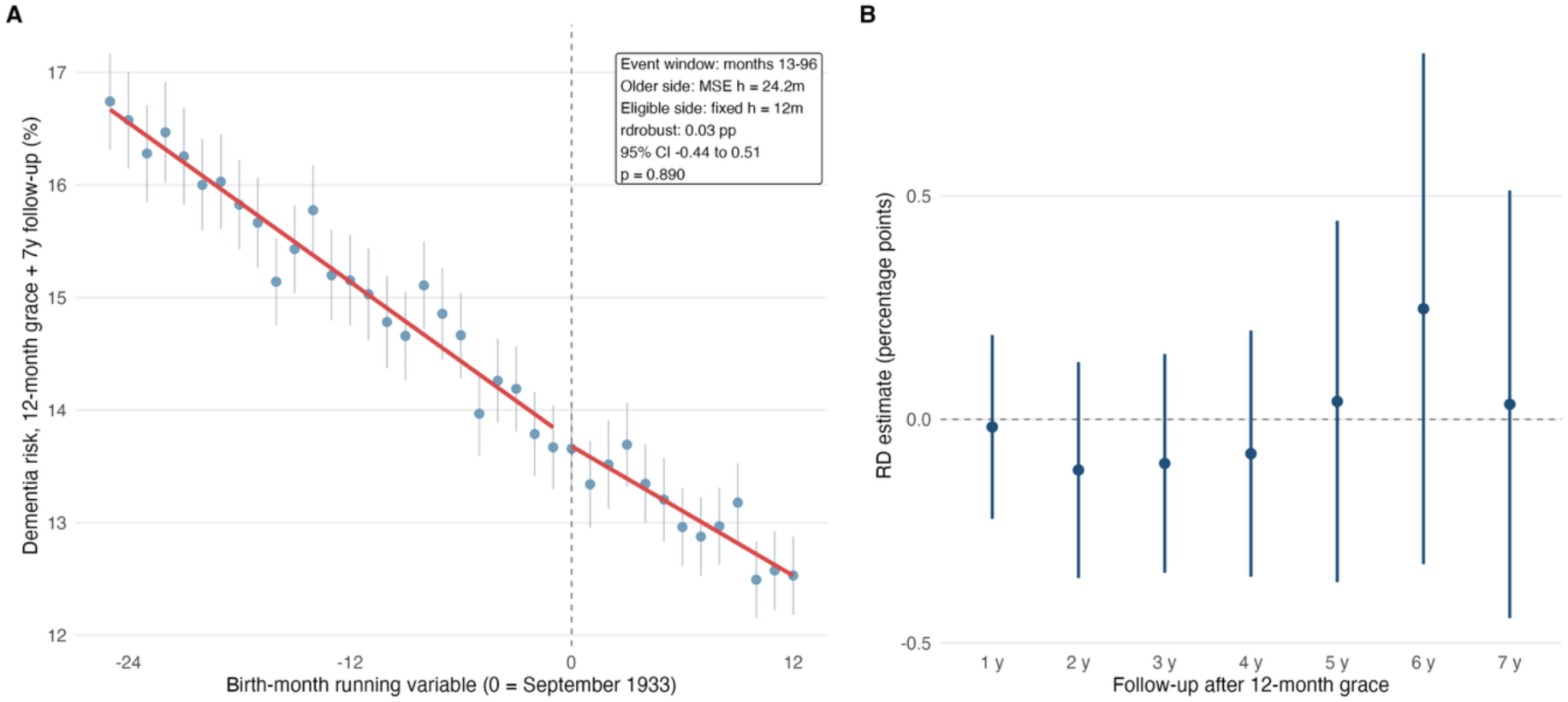
Early-follow-up dementia analysis aligned to first-year vaccine rollout. Panel A shows monthly dementia risk after a 12-month grace period and 7 years of post-grace follow-up, with the eligible side restricted to a fixed 12-month bandwidth and the older side using the MSE-selected bandwidth. Panel B shows RD estimates for 1 to 7 years of follow-up after the 12-month grace period. Estimates use HES denominators and nearest-neighbour rdrobust inference.

### Balance checks and negative controls show the importance of birth-month structure

We then went on to perform balance checks: these test whether the population either side of the RDD is similar in terms of previous diagnoses, analogous to a baseline covariate table in an RCT. We performed these for shingles, dementia and the top 20 ICD-10 codes in our dataset. These were chosen simply by frequency. For our primary HES sample we only have approximately 18 months of hospital data before the 1 September 2013 programme start, so these reflect only a small proportion of true cases and only those recorded in hospital data.

There was no evidence of discontinuity in prior dementia diagnoses (**Supplementary Figure 7**; raw p = 0.75), although there was nominal evidence of discontinuity in prior shingles diagnoses (raw p = 0.049, **Supplementary Figure 7**). Across the top 20 ICD-10 codes (**Supplementary Figure 8**; **Supplementary Table 3**), type 2 diabetes mellitus (ICD-10 E11) showed the clearest pre-index imbalance, with an estimated 0.506 percentage-point absolute difference across the cutoff (95% CI 0.17 to 0.84, p = 0.003), and personal history of malignant neoplasm (Z85) also showed a smaller nominal imbalance. We then tested the same top 20 codes as post-index negative controls using the same 8-year follow-up as the primary outcomes (**Supplementary Table 4**). Type 2 diabetes was again visually prominent (0.495 percentage points, 95% CI 0.16 to 0.83; p = 0.004), while history of malignant neoplasm did not show clear post-index imbalance (**Supplementary Figure 9**).

Visual inspection suggested that the diabetes pattern was dominated by strong month-of-birth structure rather than a sharply localized discontinuity (**Figure 5A-B**). We then formally tested whether adding birth month improved model fit beyond a smooth age trend in the whole-period series for each outcome (see **Methods** for details). This showed extremely strong birth-month structure for diabetes (p = 9.5 × 10^-51^; **Figure 5F**, **Supplementary Table 5**).

**Figure 5:**
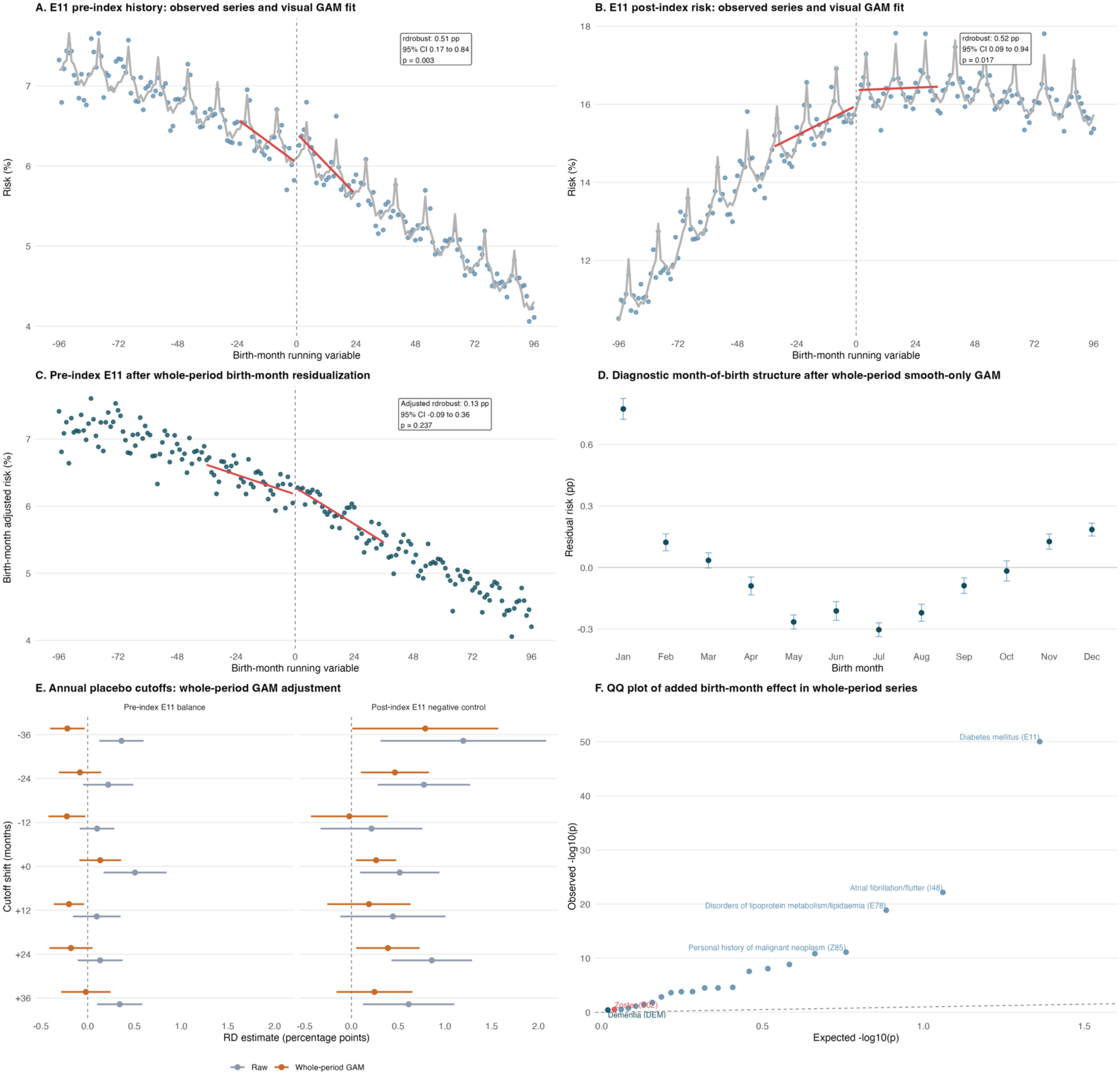
Birth-month structure as a likely explanation for the apparent type 2 diabetes mellitus (E11) pattern. Panels A and B show the observed pre-index and post-index E11 series with a visualization GAM including a smooth age/DOB term and birth month. Panel C shows the pre-index type 2 diabetes series after whole-period birth-month residualization. Panel D shows diagnostic month-of-birth structure after removing only a smooth age/DOB term. Panel E compares annual placebo-cutoff RD estimates before and after whole-period GAM-based birth-month residualization. Panel F shows a QQ plot of the added birth-month effect across outcomes using denominator-weighted GAM F tests in the whole-period series.

We then tested whether removing this birth-month seasonal component altered the RD estimates. Using all type 2 diabetes events to estimate the birth-month component, we fitted a generalised additive model (GAM) with a smooth age term and birth month as a factor (see **Methods**), then residualized out only the birth-month component before rerunning the RD analysis. This left the broad age trends intact but attenuated the pre-index type 2 diabetes imbalance from 0.506 percentage points (95% CI 0.17 to 0.84; p = 0.003) to 0.135 percentage points (95% CI -0.08 to 0.36; p = 0.24). The post-index type 2 diabetes negative-control estimate was also reduced, from 0.516 percentage points (95% CI 0.092 to 0.94; p = 0.017) to 0.265 percentage points (95% CI 0.05 to 0.48; p = 0.016). A pre-index-only GAM sensitivity gave the same broad pattern (**Supplementary Table 6**). More broadly, birth-month structure was present for several frequent hospital-coded outcomes (**Figure 5F**).

For completeness, we re-ran the primary shingles and dementia analyses using whole-period GAM-based birth-month residualization, despite the lack of evidence of birth-month structure. The primary conclusions were unchanged: shingles remained strongly protective after adjustment (−0.11 percentage points, 95% CI -0.15 to -0.08; p = 2.7 × 10^-9^), whereas dementia after 8 years of follow-up remained null (−0.14 percentage points, 95% CI -0.45 to 0.18; p = 0.40, **Supplementary Table 7**).

In sum, the negative-control analyses show that birth-month structure is a real nuisance feature of these data and can generate apparent discontinuities, particularly for diabetes, and once accounted for, the largest negative-control imbalance was attenuated, although the residual type 2 diabetes signal means this remains a useful cautionary diagnostic rather than evidence of a vaccine effect. One limitation of these analyses is that they are based on only approximately 18 months of pre-programme hospital data and therefore reflect only a small proportion of true cases and only those recorded in hospital data; absolute rates therefore do not reflect true absolute prevalence. To assess the potential impact of this limitation and to provide confidence that our generally null results were not driven by our sampling frame, we repeated balance checks using a separate set of HES records held by the Small Area Health Statistics Unit between 1 September 2003 and 31 August 2013. This longer pre-treatment lookback did not materially alter the overall pattern of results (**Supplementary Figure 10**).

### SAIL analyses do not support HES-specific bias as an explanation for the discrepant results

Our results contrast with the Welsh SAIL analysis,⁵ which shared the same vaccination policy, healthcare system, and coding, and is broadly sociodemographically similar to England. To understand why, we accessed the SAIL Databank and drew on independently conducted analyses of a separately held SAHSU HES extract with a longer lookback. We tested the three most plausible ways HES might generate a different result: (i) incomplete exclusion of prevalent dementia through lack of primary care data; (ii) a sampling frame reliant on hospital contact; and (iii) reliance on secondary care coding of dementia. Using Eyting et al.’s analytical code applied to SAIL, we reproduced a dementia ITT effect very close to the published Welsh analysis. In the closest all-source, all-sex, month-of-birth specification using full SAIL quality control, the intention-to treat estimate was -1.30 percentage points (95% CI -2.71 to -0.19, p = 0.024), compared to the reported effect of -1.30 (95% CI, -2.7 to -0.2, p = 0.022).^5^ We could then impose HES-like restrictions singly and in combination and observe whether the effect was abolished or reduced. For each candidate bias we give a theoretical argument for when it should matter and a direct empirical test in SAIL; the full set of estimates across the resulting specification matrix is shown in **Figure 6**.

**Figure 6:**
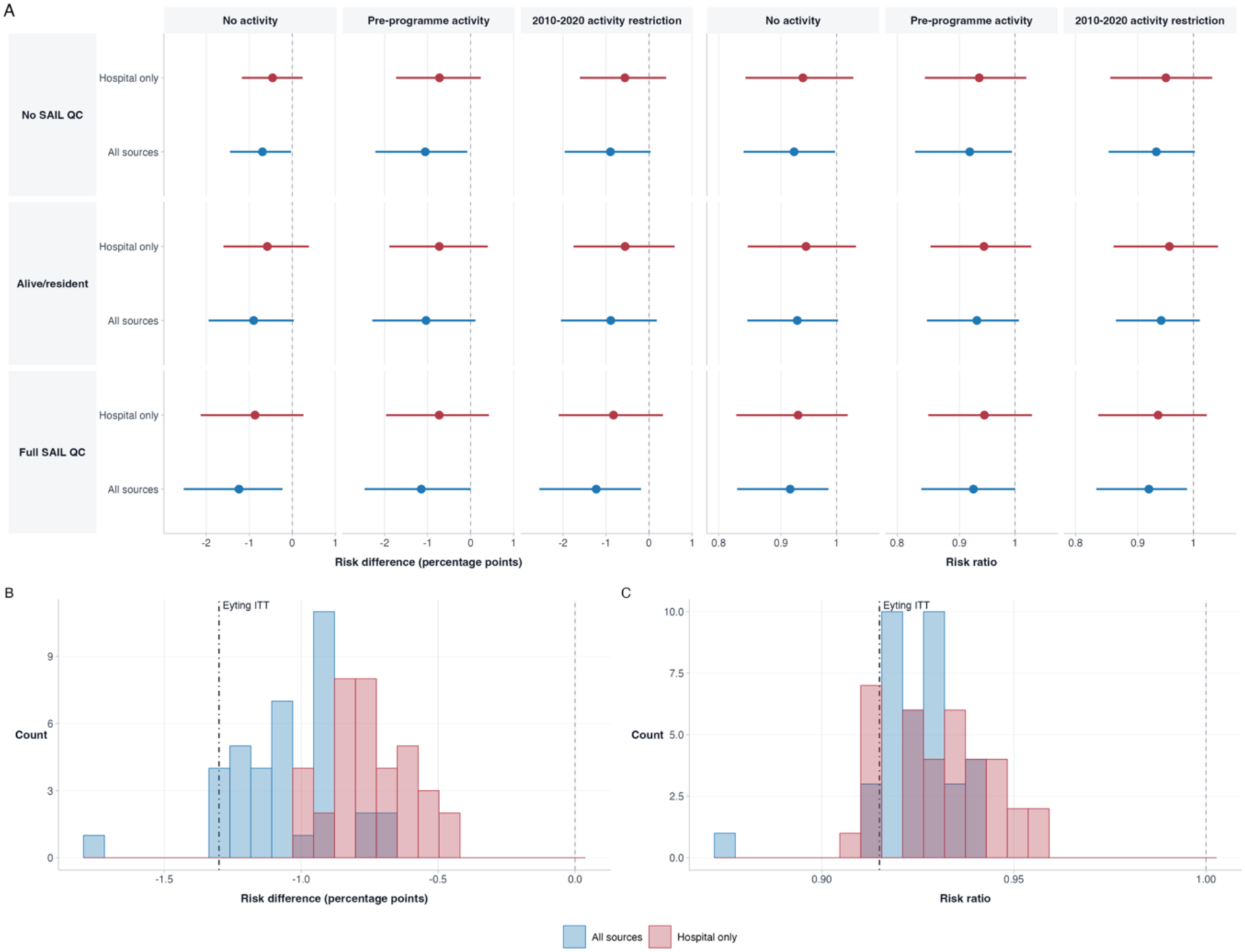
SAIL intention-to-treat (ITT) dementia estimates are stable across specification and, on the relative scale, unaffected by ascertainment. Panel A shows all-sex ITT estimates with prevalent dementia included. Rows vary SAIL quality control (no QC, alive/resident, full QC), each row shows all-source and hospital-only ascertainment, and columns vary the activity restriction used to approximate the HES sampling frame, on both the risk-difference (percentage points) and risk-ratio scales. Panels B and C show histograms of all all-sex SAIL ITT dementia estimates in the exported matrix on the absolute (risk difference) and relative (risk ratio) scales, coloured by ascertainment source; the dot-dash line marks the Eyting et al. reported ITT estimate.

### Prevalent dementia is unlikely to bias the estimate materially

One explanation for the higher dementia rate in HES than in SAIL is incomplete exclusion of prevalent disease. We excluded 2.4% of the HES population with a hospital dementia code in the 17 months before the cutoff, against 4.7% in SAIL. This is unlikely to bias the RD estimate. The continuity assumption requires only that prevalent dementia be balanced across the cutoff, and because prevalence is defined on pre-programme events, no plausible mechanism produces a discontinuous jump at the September 1933 threshold; the expected bias is also small because prevalent cases are rare (formalised in **Supplementary Note 2**). Empirically, including versus excluding prevalent cases in SAIL left the estimate essentially unchanged (ITT with prevalent cases -1.24 percentage points, 95% CI -2.52 to -0.22, p = 0.019; ITT without prevalent cases -1.30 percentage points, 95% CI -2.71 to -0.19, p = 0.024), and in the independently conducted SAHSU analysis, extending the HES lookback to 10 years, which raised exclusions from 2.4% to 2.7%, left the month-of-birth discontinuity close to zero (−0.15 percentage points, 95% CI -0.59 to 0.29, p = 0.50), as did the week-of-birth estimate (−0.16 percentage points, 95% CI -0.55 to 0.23, p = 0.42; **Supplementary Figures 11 and 12**).

### Sampling frame, denominator, and ascertainment change the scale but not the signal

SAIL includes a near-complete population register with death and migration linkage, so Eyting et al. could restrict to individuals alive and resident at programme start. This quality control changes the denominator, and therefore the absolute scale of the estimate, but not the strength of the signal, so it cannot generate a null unless the restriction is unbalanced across the threshold, for which there is no evidence. In the prevalent-dementia-excluded all-source specification, the sample size changed from 506,010 before QC to 291,524 after full QC, while the ITT estimate changed from -0.79 percentage points (95% CI -1.66 to -0.05, p = 0.037) to -1.30 percentage points (95% CI - 2.71 to -0.19, p = 0.024). In the prevalent-dementia-included matrix plotted in **Figure 6A**, requiring healthcare activity during 2010-2020 (e.g. restricting the included participants to those who might be observed in HES) reproduced the primary result (ITT -1.23 percentage points, 95% CI -2.55 to -0.18, p = 0.023). Restricting ascertainment to hospital-only coding attenuated the absolute effect (ITT -0.87 percentage points, 95% CI -2.13 to 0.26, p = 0.126), as predicted for non-differential misclassification given a hospital dementia sensitivity of about 78%, but did not reduce the relative effect.^24^

### Specification choice does not materially alter the effect on the relative scale

Running the full combinatorial matrix of these choices (QC level, sampling frame, ascertainment source, and prevalent-dementia handling) confirms the pattern (**Figure 6A**; **Supplementary Figure 13**): across the specification matrix a clear all-source dementia effect remains in SAIL, and the only material difference is the expected shift of the absolute effect toward zero under hospital-only coding (**Figure 6B**). This attenuation disappears on the relative scale (**Figure 6C**), where risk ratios cluster around 0.91 to 0.95 and are essentially unaffected by ascertainment source or specification, exactly as predicted for non-differential misclassification. The difference between our null English HES estimate and the Welsh result is therefore not explained by the sampling frame, the denominator, or reliance on hospital coding. The same matrix excluding prevalent dementia is shown in **Supplementary Figure 13.** Sex-specific versions including and excluding prevalent dementia are shown in **Supplementary Figures 14 and 15**, respectively.

The independently conducted SAHSU analyses produced similarly null estimates despite the longer pre-programme lookback and more granular week-of-birth running variable, reducing the likelihood that either feature explains the primary result (**Supplementary Figures 11 and 12**). In summary, no candidate feature of HES, alone or in combination, could reduce a Welsh-magnitude effect to our observed null, and we found no evidence that introduction of the live shingles vaccine reduced hospital-coded dementia in England.

## Discussion

We performed an RDD analysis of the September 2013 live shingles vaccination programme in England using Hospital Episode Statistics, covering 6.3 million individuals born within eight years of the eligibility threshold. We identified a clear protective effect on hospital-coded shingles across the main analyses. Despite a roughly 18-fold increase in the sample size compared with the Welsh SAIL analyses reported by Eyting et al.^5^, we did not find evidence of a corresponding effect on hospital-coded dementia. Extensive sensitivity analyses were broadly null, with the only nominal deviation being a short-term follow-up signal that attenuated to the null by two years. The credibility of our null result rests on whether features of HES that differ from SAIL could plausibly have attenuated a Welsh-magnitude effect to our observed result. We addressed this by accessing SAIL, which (i) previously showed the effect, (ii) has a different sampling frame, and (iii) includes primary care and death register data as well as hospital data. As set out in the **Results**, for each of the three candidate biases (prevalent dementia, sampling frame and denominator, and secondary care ascertainment), theoretical and empirical lines of evidence do not support these generating a large bias in our estimates.

The major strength of our study is its very large HES-derived hospital-activity cohort from England, which is considerably larger than the Australian^8^ and Ontario^7^ datasets examining this question. A further strength is the independent analysis of a separately held, aggregated HES extract by the SAHSU team. The two extracts had complementary strengths: the primary extract permitted person-level cohort construction and included inpatient and outpatient activity, whereas the SAHSU extract provided a longer pre-programme lookback and week-of-birth resolution. The concordant null estimates across extracts, analytical teams, and running-variable resolutions reduce the likelihood that the result reflects extract-specific construction or analytical implementation.

The major limitations of our study relate to the nature of our dataset. We do not have access to primary care data, death data, or prescription data. Primary care and vaccination data would improve dementia ascertainment and allow complier-adjusted analyses. Linked death data would be valuable because death is a competing event for hospital-coded dementia: individuals who die before receiving a hospital dementia code can no longer contribute a subsequent first hospital-coded dementia event. Therefore, death can influence the observed cumulative incidence primarily through the numerator, by changing who remains alive and observable for hospital-coded dementia, rather than by generating a discontinuity in the denominator; such competing-risk effects would be most concerning if they changed discontinuously at the eligibility threshold.

The strongest statistical evidence for a signal was at early follow up (1 year). We do not think this is a plausible signal for several reasons. Firstly, the known delay in hospital-recorded diagnosis means that many people admitted in the first year of follow up will have dementia that actually was diagnosed in primary care before the index date. Secondly, the delayed rollout is documented in official programme data^23^. **Supplementary Figure 5** separately shows that some dementia diagnoses preceded vaccination. Assuming the vaccine does not work instantly, it is therefore highly implausible that outcomes occurring in the first year after introduction of the vaccine are related to the vaccine and are more plausibly related to the play of chance.

Several considerations make a large HES-specific competing-risk artefact unlikely. Hospital contact before death is common in this age group: in England, 81% of people aged 75 years or older who died in 2017 had at least one hospital admission in their final year of life^25^ (and are therefore likely observed), and contemporary ONS-HES linked end-of-life data show that 51% of people dying of dementia had an inpatient admission even within the final 6 months of life.^24^ In addition, any bias capable of explaining the null HES estimate would need to operate discontinuously at the September 1933 eligibility threshold, whereas our HES and ONS denominators were highly correlated across month-of-birth bins and showed no visible discontinuity at the threshold. We therefore cannot exclude competing-risk effects, particularly at longer follow-up, but these are unlikely on their own to explain a large discrepancy between HES and SAIL estimates. We should also note the difference in magnitude between our estimates from HES and from the ONS. Although these may reflect deduplication issues and transient populations, the large difference between estimates means we should be careful about the absolute scale of the observed effect which relies on the denominator.

Importantly, this means our estimates should not be treated as population level effects and the absolute size of the effect can differ based on the denominators. For example, our primary estimate on dementia in hospital varies from around 0.06pp to around 0.15pp when using HES data from two different sources, reflecting both noise around the point estimate but also differing denominators. However, as we do not identify a robust causal effect, and do not think denominator choice leads to bias, these are less relevant in interpretation.

The discrepancy between estimates from this RD analyses and the previous analyses in Australia, Wales, and Ontario is marked. For the Wales analyses, the upper end of our England estimate (0.4 percentage point decrease) is within the lower end (0.2 percentage point decrease) of the published Welsh estimate.^5^ It is therefore conceivable that these estimates are concordant. However, for the Ontario and Australia analyses, the reported effects are much larger and cannot easily be reconciled with our result. One potential explanation for the difference between these estimates is that the potential researcher degrees of freedom in choosing how to analyse RD designs is larger than in randomised trials, with choices over bandwidth, estimator, estimator specification, alongside numerous other choices around data source and exclusion and inclusion criteria. This means that estimates are necessarily more variable than those identified in ITT analyses of RCTs, and visual evidence of this can be seen in **Figure 6**, where changes in the data source lead to variation in the reported estimate. Note, these analyses all used the same estimation strategy (rdrobust, with settings from Eyting et al), so are an underestimate of variability in RD estimates. This is not to suggest these analyses are meaningless, but that the reported confidence intervals likely underestimate the true variability driven by analytical choices.^25^

Where does this leave us with respect to the potential for shingles vaccination preventing dementia? Firstly, we must be clear that this evidence relates entirely to the live vaccine, which is now not widely used, having been largely replaced by the recombinant shingles vaccine. Some evidence^11^ using a weaker quasi-experimental design than the date-of-birth eligibility cutoff analyses used to study the effects of the live-attenuated shingles vaccine, suggests a protective effect of the recombinant shingles vaccine (or perhaps the adjuvant in the recombinant vaccine^12^) on dementia-related outcomes. The evidence we present here does not support a large effect of live shingles vaccine eligibility on hospital-coded dementia in England. However, it does not by itself settle the wider question across settings, data sources, or vaccine formulations. This result may reflect a true null effect in this setting, residual differences between datasets or analytic choices, or sampling variability in a still developing literature.

In our view, the English HES-based evidence presented here is more compatible with no large effect on dementia than with a strong protective effect, but the wider epidemiological picture remains mixed and still merits further investigation, given the limitations of hospital-based coding. Further epidemiological investigations should be performed using data from the whole of England including primary care data (e.g. using OpenSAFELY, CPRD, or other related datasets). Additionally, similar investigations of the recombinant shingles vaccine should be performed, where many countries switched rapidly from the live to recombinant vaccine. These are less likely to be biased in single-payer systems like the UK. Other epidemiological approaches like genome wide association studies and Mendelian randomisation should be considered, accepting the challenges of genetic epidemiology in infection.^27^ Finally, randomized controlled trials (RCT) should be performed. Two large, pragmatic RCTs in Finland (NCT07502560) and Denmark (NCT07485283) with dementia as an outcome are planned although given the timeline to follow up this may take a number of years to read out.

## Conclusion

In a large English hospital dataset, we found no meaningful evidence that eligibility for shingles vaccination reduced hospital-coded dementia.

## Methods

### Study design and policy threshold

We used a regression discontinuity design (RDD) centred on the September 1933 age-eligibility threshold for the live shingles vaccination programme in England. Under the exact date-of-birth rule, the eligibility threshold fell within the September 1933 birth month: individuals born before the threshold were ineligible and those born on or after it were eligible. Because only month and year of birth were available under our HES data access agreement, the running variable was defined as month of birth relative to September 1933. In the primary RD analyses, the September 1933 birth-month bin was assigned to the eligible side of the cutoff, consistent with the policy threshold falling within that month and with most people born in September 1933 being eligible. The small ineligible fraction of this boundary month could not be separated at day level. Analyses specifically examining birth-month structure excluded this boundary month because exact day of birth was unavailable.

### Data sources

We used Hospital Episode Statistics (HES), accessed through the University of Bristol under NHS Digital Data Access Request Service agreement DARS-NIC-17875-X7K1V. HES is an administrative dataset of NHS inpatient and outpatient activity in England with ICD-10-coded diagnoses (up to 20 per event) and person-level linkage via an anonymised NHS identifier. HES does not include individual-level vaccination records, linked primary care data, or complete death-register ascertainment; our estimates are therefore intention-to-treat effects of vaccine eligibility on hospital-coded outcomes. HES annual extracts follow the UK financial year; these data covered outpatient and inpatient hospital activity in England from 1 April 2012 to 31 March 2022. The person-level records allowed outcome-specific exclusions and identification of first post-index events before aggregation into birth-month-by-sex cells for RD estimation.

In parallel, the SAHSU team independently analysed HES data held under a distinct data-access agreement (DARS-NIC-204903-P1J7Q-v7), covering inpatient healthcare events in England from 1 September 2003 to 31 August 2023 (detailed in **Supplementary Information)**. The SAHSU extract available for analysis comprised aggregated week-of-birth cohorts and additionally included linked HES-Office for National Statistics (ONS) mortality data.

We accessed SAIL Databank data through Cardiff University within the SAIL Gateway secure research environment. The study was approved by the SAIL Information Governance Review Panel (application number 0998). The SAIL analyses used linked demographic (Welsh Demographic Services Database; WDSD), hospital (Patient Episode Database for Wales; PEDW), primary-care (Welsh Longitudinal General Practitioner dataset; WLGP), mortality (WDSD and Annual District Death Extract; ADDE), and vaccination-timing data (contained within WLGP) to reproduce the Eyting et al. specifications and to construct HES-like restrictions.

### Cohort construction and denominators in HES

The analytic cohort comprised individuals born within 96 months of September 1933 with any recorded HES activity across the study window. Denominators were stratified by month of birth and sex. The default denominator was the HES hospital-activity denominator for each birth-month/sex cell. For each primary outcome, we defined a non-prevalent cohort by excluding individuals with a prior hospital-coded diagnosis of the outcome before 1 September 2013.

As a secondary denominator, we used ONS-derived resident-population estimates constructed from 2011 Census CT1315 date-of-birth counts by sex and updated using cumulative all-cause mortality counts around the programme period. Where exact date of birth is unavailable in the Census, ONS assigns the first day of the month; the resulting first-of-month excess was redistributed across other days in the same month while preserving monthly totals. The ONS-denominated analyses used full ONS resident-population counts. Because the aggregate ONS data could not be restricted by the person-level prior-diagnosis exclusions applied in HES, these analyses combined the non-prevalent HES outcome numerator with the full ONS denominator. They should therefore be interpreted as denominator-sensitivity analyses rather than estimates in exactly the same non-prevalent cohort as the primary HES analysis. For this reason and because denominator, outcome ascertainment, and recorded date of birth were drawn from HES, we treated the ONS series as a sensitivity denominator and used HES denominators as primary (**Supplementary Note 1** provides further detail). Code to generate our ONS denominator is available in the code availability section.

For our primary analyses on dementia and shingles, we further restricted the denominator by excluding prevalent cases, by excluding any participant who had a record of the relevant disease prior to the index period. As our data only has a lookback of around 18 months, this will miss a proportion of prior cases.

### Outcome definitions in HES

In HES, the primary positive-control outcome was first hospital-coded shingles (ICD-10 B02) in any diagnosis position after 1 September 2013. The primary dementia outcome was first hospital-coded dementia using the Eyting et al. codelist^5^, with a narrower alternative dementia definition used as a sensitivity analysis.

Hospital diagnosis codes were cleaned by removing non-alphanumeric characters and converting to uppercase; shingles and negative-control outcomes were defined using three-character ICD-10 roots, thereby including all subcodes under each root, while dementia was defined using the prespecified ICD-10 codelist, with an alternative dementia definition based on codes beginning F00, F01, F03, F051, or G30. Full ICD-10 definitions are given in **Supplementary Table 8**. All outcomes are hospital-coded; primary care and death certificate data were not available.

### Follow-up and outcome counting in HES

Post-index events were counted from 1 September 2013 over a fixed follow-up window, with 8 years as the primary horizon. The maximum post-index follow-up available in the extract therefore ran through to 31 August 2021. Shorter follow-up windows from 1 to 7 years were treated as sensitivity analyses. For pre-index balance analyses, we counted hospital-coded diagnoses observed in the available HES lookback period (approximately 18 months). These pre-index counts therefore reflect recent hospital-coded history rather than lifetime disease prevalence. The independently conducted SAHSU analyses used an approximately 10-year pre-programme lookback and a week-of-birth running variable.

### Primary regression discontinuity analysis

Our aim was to match the Eyting et al. analysis closely, recognizing the differences in datasets which necessitate small changes in approach. Individual-level data were aggregated into counts of first events and total population by month-of-birth bin and sex. RDD estimates were obtained using rdrobust^19^, with the event proportion as the outcome and denominator counts as analytic weights. We aimed to match the published Eyting et al. rdrobust implementation where possible: local polynomial RD centred at the eligibility cutoff, a triangular kernel, data-driven bandwidth selection, nearest-neighbour variance estimation, and mass-point adjustment for the discrete running variable.

We made two explicit changes in applying rdrobust to these HES data compared to Eyting et al.^5^ First, unlike the Eyting et al. code, which combined the conventional coefficient with robust confidence intervals and p-values, we consistently report the robust bias-corrected coefficient, confidence interval, and p-value returned by rdrobust. Second, we used nearest-neighbour variance estimation with three matches, the rdrobust default, because it is the standard RD-specific variance estimator and estimates local outcome variation using neighbouring values of the running variable.^19^

We include the sandwich estimator (“HC0”) used in the prior paper as a sensitivity analysis. Estimates are reported as absolute percentage-point differences at the threshold. Because individual vaccine uptake is not observed in HES, we report intention-to-treat effects of eligibility and do not estimate fuzzy RDD or complier-average causal effects.

### Sensitivity analyses

Our further sensitivity analyses comprised: (i) varying follow-up from 1 to 8 years; (ii) grace periods of 0 to 12 months before outcome counting begins; (iii) restricting outcomes by diagnosis position (any, 1-2, other); (iv) fixed bandwidths (h = 12 and h = 24 months); (v) HC0 plug-in heteroskedasticity-robust variance estimation instead of nearest-neighbour variance estimation; (vi) local quadratic rdrobust fits (p = 2, q = 3); (vii) RDHonest as an alternative estimator under honest inference; (viii) annual placebo cutoffs at ±12, ±24, and ±36 months from the true September 1933 threshold; (ix) ONS rather than HES denominators; (x) excluding the September 1933 boundary month; and (xi) a longer pre-programme lookback period of 10 years to identify prevalent dementia using the SAHSU HES extract.

Together these test whether the main estimates are sensitive to ascertainment window, biological lead time, diagnostic coding priority, bandwidth choice, local functional form, threshold identification, denominator construction or incomplete exclusion of prevalent dementia cases due to the restricted lookback period.

Additionally, to specifically focus on the plausibility of signals at early follow-up, we performed an analysis with a fixed 12-month eligible-side bandwidth, an MSE-selected older-side bandwidth, and a 12-month grace period before outcome counting. This was chosen to ensure that outcomes were plausibly post-vaccination and downstream of vaccination. Official programme data show that vaccination in the first year was offered to the routine 70-year-old and catch-up 79-year-old cohorts, with most doses administered in the first few months and coverage in both cohorts exceeding 45% by the end of January 2014.^23^ A 12-month grace period was used assuming the vaccine plausibly takes at least 6 months to reduce hospital admission with dementia, which would still be an extremely fast effect.

Separately, because the only nominal dementia association in the primary analysis occurred at 1 year, we repeated annual placebo-cutoff analyses for the 1-year endpoint at ±12, ±24, and ±36 months from the true cutoff.

### Negative controls and balance outcomes

To test whether any apparent discontinuity reflected broader coding or cohort structure rather than vaccine eligibility, we constructed a deterministic set of negative-control outcomes. After excluding dementia codes and shingles, we selected the 20 most frequent three-character ICD-10 roots over the post-index period. These were analysed with the exact same RDD framework both pre- and post-index; post-index negative-control analyses used the same 8-year follow-up window as the primary post-index outcomes.

### Birth-month structure

Because the running variable is defined in month-of-birth bins rather than exact date of birth, birth-month structure could create apparent local discontinuities unrelated to vaccine eligibility. We treated this as nuisance structure rather than as a causal adjustment. For each monthly outcome series, we fitted denominator-weighted generalised additive models using mgcv to the observed event proportion, with model p ∼ s(t_year, k = 12) + birth_month, where t_year is months from the September 1933 threshold divided by 12 and birth_month is a 12-level categorical factor. Models were fitted by maximum likelihood, with denominator counts as analytic weights.

For the primary E11 (diabetes mellitus) diagnosis, we summed pre-index and post-index E11 cases and denominators within each month-of-birth bin before fitting the GAM, excluding the boundary month because exact day of birth was unavailable. To residualize the series, we subtracted only the fitted birth-month component: the fitted value for the observed birth month minus the mean fitted value across all 12 birth months at the same t_year. This retained the smooth age/date-of-birth trend while removing the estimated birth-month component. We then repeated the RD analyses on the residualized outcome. A pre-index-only GAM, fitted in the pre-index E11 series and applied to the post-index series, was retained as a sensitivity analysis for E11. For primary shingles and dementia residualization checks, we fitted analogous whole-period GAMs using the corresponding pre-index and post-index outcome series; these primary-outcome checks retained the boundary month to mirror the primary RD analyses. We tested the contribution of birth month by comparing denominator-weighted GAMs with and without the birth_month term using F tests: p ∼ s(t_year, k = 12) versus p ∼ s(t_year, k = 12) + birth_month.

### SAIL analyses

In SAIL, we adapted the RD code published by Eyting et al. (available at^27^) to take different parameters in analysis and to use newer iterations of the same tables for each data source. We ran the code and then varied, singly and in combination, all-source versus hospital-only dementia ascertainment; inclusion versus exclusion of prevalent dementia; no SAIL quality control, alive/resident-only quality control (requiring participants to be alive and resident in Wales at the index date), and full quality control (which is equivalent to the original Eyting et al. quality control in SAIL); no hospital activity restriction, restriction to those with recorded hospital (in-patient or out-patient) activity before programme start, and restriction to individuals with recorded hospital activity during 2010-2020; month versus week running variables; and prespecified bandwidth and grace-period variants.

### Software and reproducibility

We used R v4.6.0 for secure data processing, downstream statistical analysis, and figure generation. The secure export stage used DuckDB and data.table to construct summary cells from the source HES. The main analyses used rdrobust and RDHonest, and the birth-month structure analyses used mgcv.

The source HES data are not shareable by the authors in line with NHS England policies. Access is governed through the relevant NHS data-access processes and University of Bristol/Small Area Health Statistics Unit policies. R code for our primary analyses is available at github.com/gushamilton/HES_zoster. R code for other analyses will be available on publication.

### Ethical approval

We were provided with routinely collected Hospital Episode Statistics data under licence from NHS Digital (DARS-NIC-17875-X7K1V). The licence allows us to use the information under Section 261 of the Health and Social Care Act 2012, 2(b)(ii): “after taking into account the public interest as well as the interests of the relevant person, considers that it is appropriate for the information to be disseminated”.

SAIL analyses used anonymised linked data held within the SAIL Databank and were conducted under SAIL information-governance approvals in the secure SAIL Gateway environment. SAHSU holds approvals both from the London - South East Research Ethics Committee (22/LO/0256) and from the Health Research Authority - Confidentiality Advisory Group (20/CAG/0028).

### Funding and support

FH’s time was funded by the NIHR Clinical Lectureship Programme and an MRC Clinician Scientist Fellowship (OPP844). This research was supported by the National Institute for Health and Care Research (NIHR) Applied Research Collaboration (ARC) West. SAHSU is part of the MRC Centre for Environment and Health supported by the UK Medical Research Council, Grant number: MR/L01341X/1 and Health Data Research UK (HDR UK) in the Social and Environmental Determinants of Health Driver Programme (HDRUK2023.0029), which is funded by UK Research and Innovation, the Medical Research Council, the British Heart Foundation, Cancer Research UK, the National Institute for Health and Care Research, the Economic and Social Research Council, the Engineering and Physical Sciences Research Council, Health and Care Research Wales, Health and Social Care Research and Development Division (Public Health Agency, Northern Ireland), Chief Scientist Office of the Scottish Government Health and Social Care Directorates. PE, BD and APM acknowledge infrastructure support for the Department of Epidemiology and Biostatistics provided by the NIHR Imperial Biomedical Research Centre (BRC NIHR203323). GDS received support from the National Institute for Health and Care Research (NIHR) Biomedical Research Centre: Bristol (BRC NIHR203315). DA was funded by an NIHR Advanced Fellowship award (NIHR159462). PE received support from the UK Dementia Research Institute [award number UK DRI-5001] through UK DRI Ltd, principally funded by the Medical Research Council. MBS and VEP are by the UK Dementia Research Institute, which is funded by the Medical Research Council (UK DRI-5001), Alzheimer’s Research UK, and Alzheimer’s Society. MDC acknowledges financial support from the National Science Foundation through grant SES-22415755 and the John Simon Guggenheim Memorial Foundation through a 2026 Guggenheim Fellowship. This work uses data provided by patients and collected by the NHS as part of their care and support. Hospital Episode Statistics data are copyright © 2026, re-used with the permission of NHS England. All rights reserved. The views expressed in this article are those of the author(s) and not necessarily those of the NHS, the NIHR, or the Department of Health and Social Care.

## Data availability

The data used in this study are not publicly available and cannot be shared by the authors because they contain routinely collected health information accessed under data-use agreements. Hospital Episode Statistics data were accessed through NHS England under agreements DARS-NIC-17875-X7K1V and DARS-NIC-204903-P1J7Q-v7. Researchers may apply for access through the relevant NHS England data-access processes, subject to the necessary approvals and agreements. SAIL data are held within the secure SAIL Gateway and may be accessed only following approval through the SAIL Databank’s standard application and information-governance processes. The authors are not permitted to redistribute either the underlying individual-level data or the restricted aggregated extracts.

