## Supplementary Methods for "Eligibility for shingles vaccination and hospital-coded dementia in England and Wales: a regression discontinuity analysis in England"

**Supplementary Information:**

**Small Area Health Statistics Unit HES sensitivity analysis**

These analyses were conducted independently from the primary analysis as the underlying HES datasets were held under different data governance and access agreements within distinct Secure Research Environments. The analyses were used to assess the potential impact of a longer pre-programme lookback to exclude prevalent dementia cases and a more granular week-of-birth running variable on the primary findings.

***Population***

We used Hospital Episode Statistics (HES) inpatient data for England between 1 September 2003 and 31 August 2023, aggregated into sex-specific week-of-birth cohorts born between 2 September 1925 and 1 September 1941 and restricted to individuals whose recorded address at the inpatient event was in England. Individuals who had died prior to programme implementation (1 September 2013) were identified using linked HES-ONS mortality data.

Our primary study denominator comprised individuals in these birth cohorts who had at least one recorded inpatient hospital healthcare event during the follow-up period (1 September 2013-31 August 2023). To construct an at-risk cohort, we excluded individuals alive on 1 September 2013 with a hospital-coded dementia diagnosis in any diagnosis position during the 10 years preceding programme implementation (1 September 2003-31 August 2013).

As a secondary study denominator, we included individuals alive on 1 September 2013 who had at least one recorded inpatient hospital healthcare event between 1 September 2003 and 31 August 2023 and no hospital-coded dementia diagnosis during the 10 years preceding programme implementation.

For pre-index balance analysis, the denominator comprised individuals with at least one recorded inpatient hospital healthcare event between 1 September 2003 and 31 August 2013.

***Outcome definitions***

As in the primary analysis, our dementia outcome was first hospital-coded dementia using the Eyting et al. codelist (F00-F03; F051; G30; G310; G311; G318; I673). Post-index events were counted from 1 September 2013 in six-monthly follow-up intervals up to eight years, the primary horizon.

For pre-index balance analyses, the number of individuals with hospital-coded diagnoses was counted between 1 September 2003 and 31 August 2013. Outcomes were examined without restricting to at-risk individuals, such that estimates reflect the proportion of individuals with at least one hospital-recorded diagnosis before vaccine implementation.

***Statistical analysis***

Regression discontinuity analyses followed the same general analytical framework as the primary analyses. Outcome risks were calculated using a week-of-birth running variable and analysed using local linear regression discontinuity models with triangular kernel weights, mean squared-error optimal bandwidth selection, and robust bias-corrected inference. Additional sensitivity analyses collapsed week-of-birth cohorts into four-week intervals to assess whether the specification of the running variable materially influenced findings.
