## Supplementary Note 1 for "Eligibility for shingles vaccination and hospital-coded dementia in England and Wales: a regression discontinuity analysis in England"

**Supplementary Note 1: Denominator choice and date-of-birth heaping**

As HES conditions on hospital exposure, it does not provide a complete population denominator. This may matter in a regression discontinuity analysis because population size varies across dates of birth. However, hospital contact is common in this age group over the study period, making either HES-derived denominators, which condition on hospital contact, or ONS resident-population denominators plausible. This note explains the rationale for using the HES denominator in the primary analysis and the limitations of the ONS sensitivity denominator.

The main analyses use HES-derived denominators, with ONS-derived denominators retained as secondary checks. The major reason for using HES as the primary denominator is that the denominator, outcome ascertainment, and recorded date of birth are all drawn from the same administrative system, which reduce the risk of bias generated by mismatch of the denominator and numerator. One additional major issue is around January months of births.

The ONS-derived denominator was constructed from 2011 Census CT1315 date-of-birth counts by sex, updated using cumulative all-cause mortality counts around the programme period. This gives an external resident-population denominator, but it has an important limitation for date-of-birth analyses: where exact date of birth is unavailable, ONS assigns the first day of the month. This creates first-of-month mass points. In our analysis, excess first-of-month counts were redistributed across other days in the same month, preserving monthly totals, but the 1 January problem cannot be fully solved because genuine 1 January births and administratively assigned 1 January births cannot be separated.

**Figure SN1** below is the clearest example of why this matters in the SAHSU week-of-birth analyses. The point estimate for eight-year hospital-coded dementia remains null using the ONS-derived denominator, but the left panel shows observations sitting systematically away from the main fitted trend (1^st^ week of January).

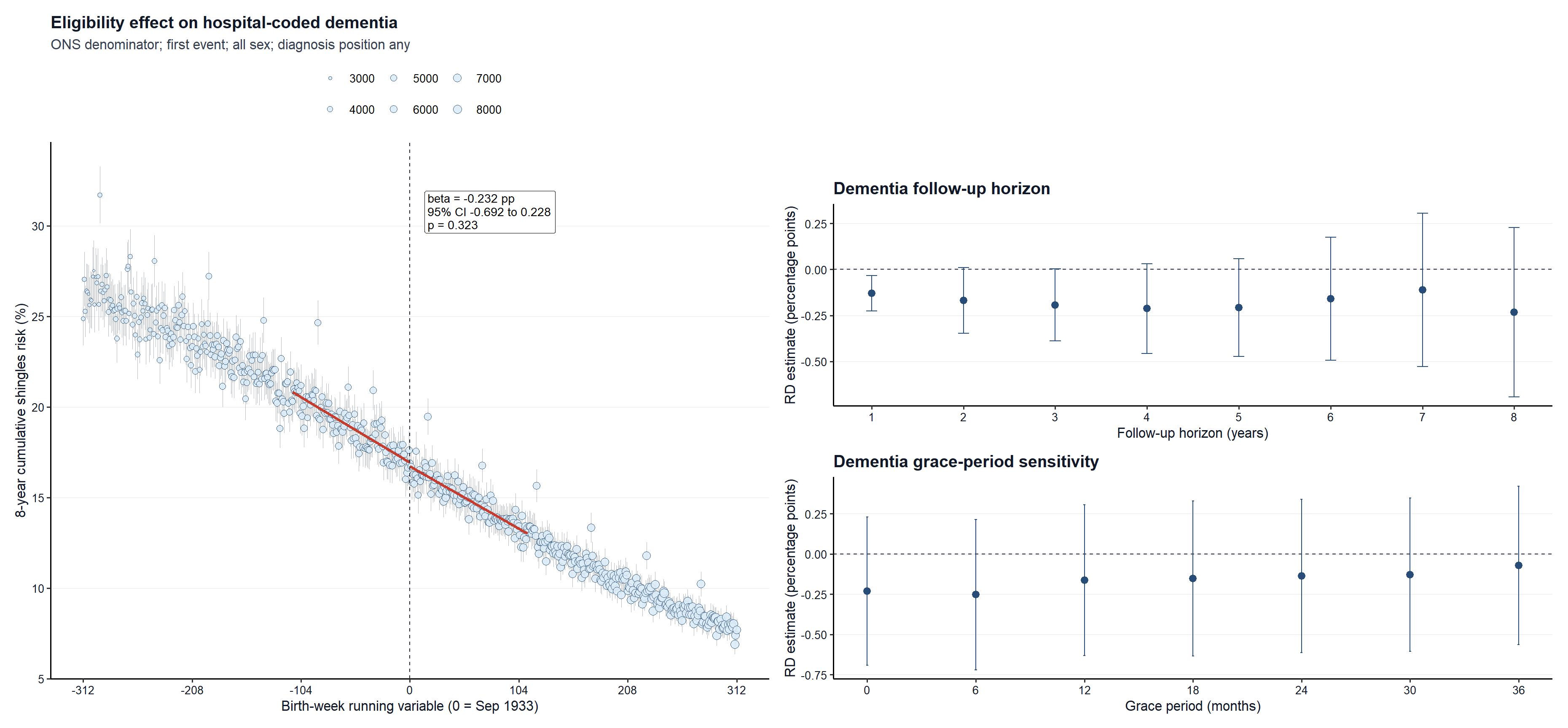

**Figure SN1.** Worked example using the ONS-derived denominator in the SAHSU all-sex week-of-birth dementia analysis. The estimate remains null, but the left panel makes the residual denominator/running-variable structure visible for the first week in January. This supports treating week-of-birth ONS-denominator analyses as sensitivity analyses rather than as the primary specification.

This can threaten RD inference because it can lead to changes in local denominator structure. The denominator defines the observed cumulative risk and contributes to the precision weights. If the denominator has local administrative structure, a smooth numerator can be turned into an outcome risk with local curvature, apparent irregularities, or altered standard errors. This is especially visible at week-of-birth resolution; month-of-birth analyses average over more of this daily structure, and is demonstrated in our Type 2 diabetes analysis in the main text.

This does not necessarily lead to bias, but it can create artificial structure. The RD design would be threatened if potential outcomes, outcome measurement, or denominator construction changed discontinuously exactly at the eligibility threshold for reasons other than vaccine eligibility. The first-of-month and 1 January artefacts are nuisance features of the running variable and administrative data systems. Provided these artefacts are smooth through the 2 September 1933 eligibility threshold, they should not themselves create a causal discontinuity. The concern is instead that they can make local trends, weights, and high-resolution sensitivity analyses less stable, and we show that this can drive artificial effects in our type 2 diabetes analyses, suggesting this is not simply a theoretical concern.

**Figures SN2** and **SN3** show further data from SASHU describing the differences between these datasets. The raw ONS-derived denominator contains repeated first-of-month spikes. Redistribution improves the series and brings it closer to HES, but residual 1 January structure remains. This is why the ONS analyses are useful as external checks, while the HES-denominated analyses remain the more defensible primary specification.

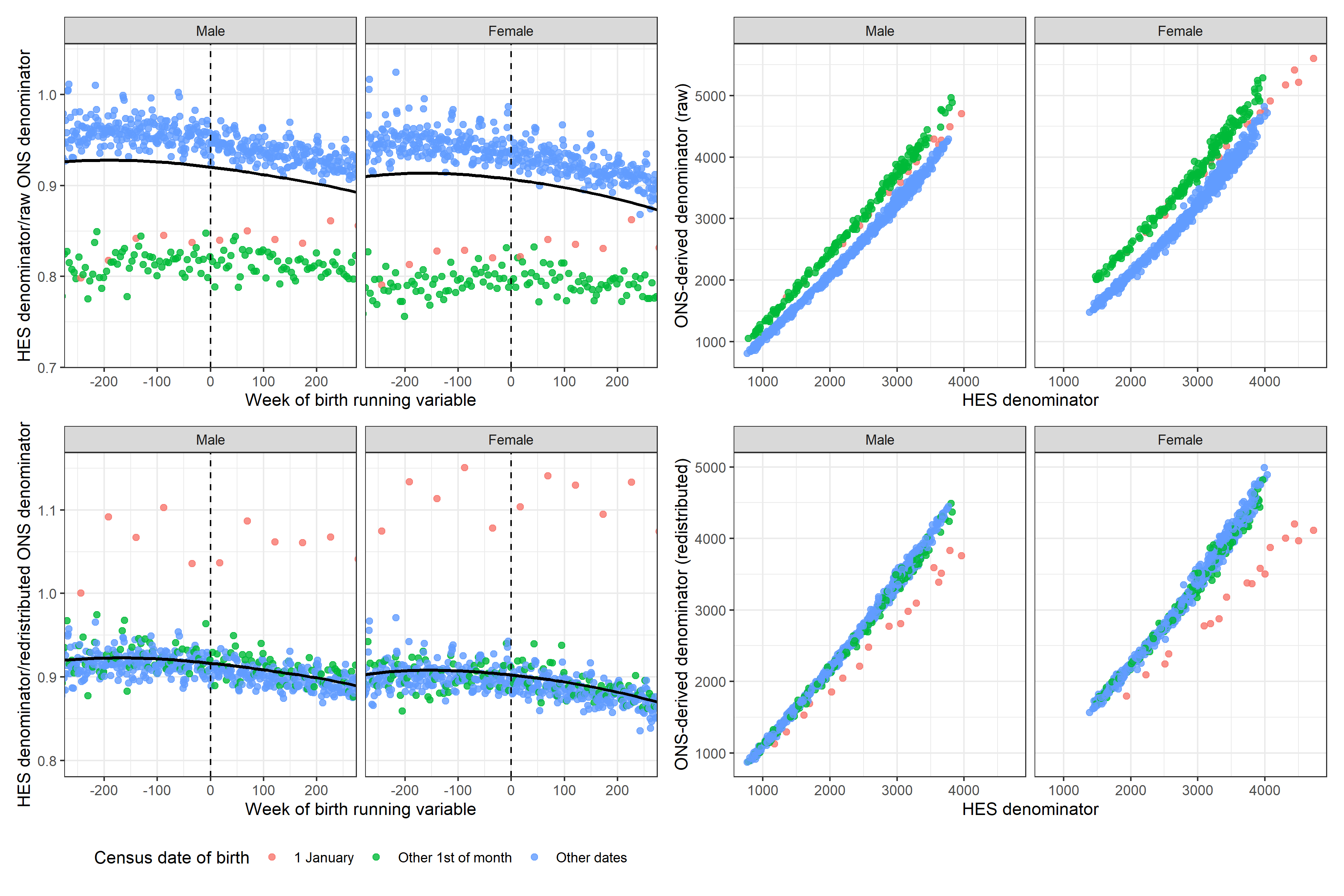

**Figure SN2.** SAHSU week-of-birth comparison of HES and ONS-derived denominators. Red points mark 1 January, green points mark other first-of-month dates, and blue points mark other dates. Redistribution improves agreement between ONS and HES, but residual 1 January structure remains.

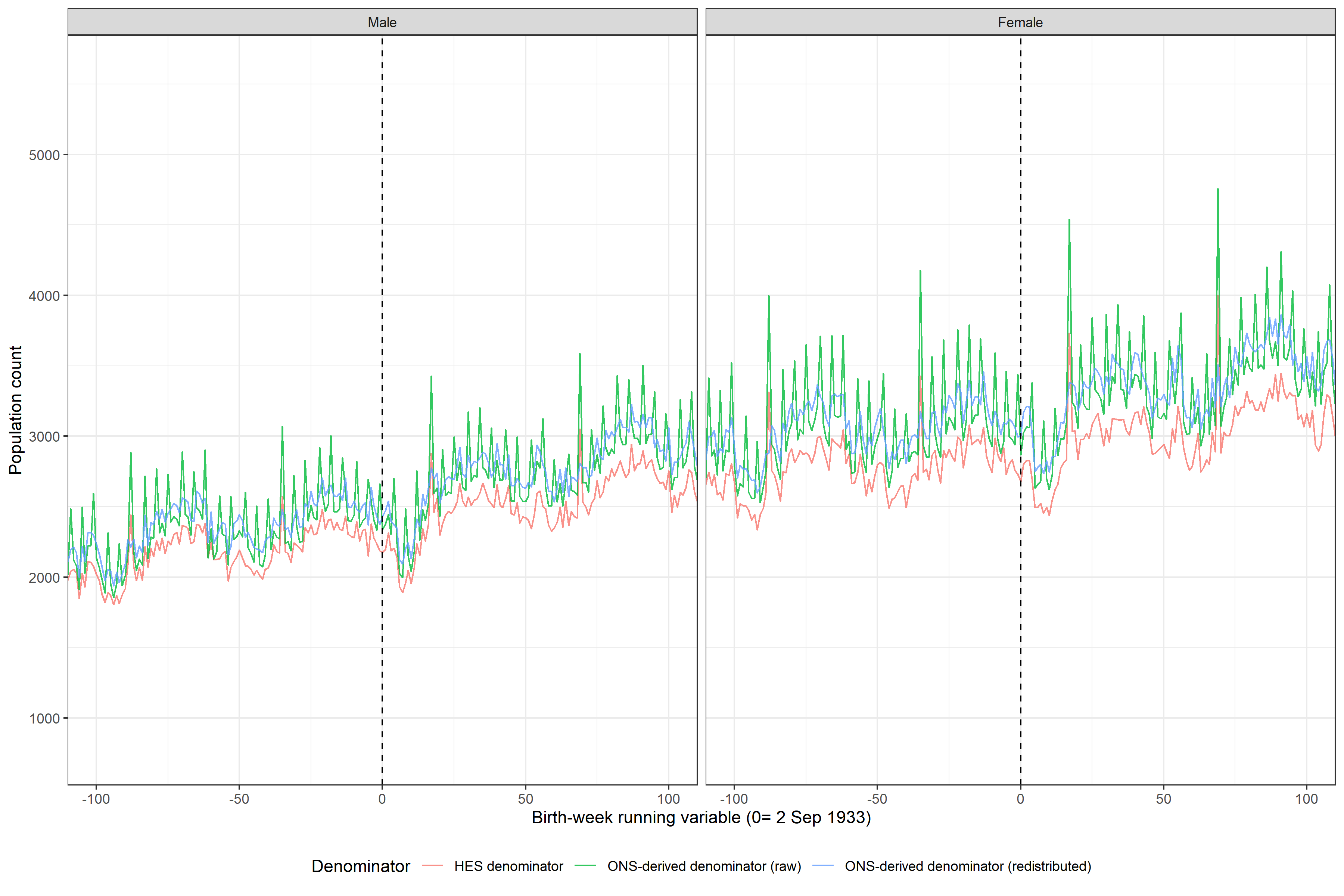

**Figure SN3.** SAHSU week-of-birth denominator series around the eligibility threshold. The raw ONS-derived denominator shows repeated first-of-month spikes; the redistributed denominator is smoother and closer to HES, but local structure remains visible. The dashed line marks the 2 September 1933 threshold.

In short, the denominator diagnostics support the use of HES denominators for the primary analyses. HES is not a clean resident-population denominator, so absolute risks should not be overinterpreted as national population risks. However, HES is internally aligned with the hospital-coded outcomes and with the recorded date of birth used to define eligibility. The ONS denominator is a useful sensitivity analysis and helps show that the null dementia result is not driven by denominator choice, but its first-of-month and 1 January structure makes it less suitable as the main high-resolution RD denominator.
