## Supplementary Note 2 for "Eligibility for shingles vaccination and hospital-coded dementia in England and Wales: a regression discontinuity analysis in England"

**Supplementary Note 2: Effect of prevalent dementia cases on RD estimates**

Let $P_{\mathrm{prev}}$ denote the probability of prevalent dementia at the threshold, that is dementia with onset before $t_{0}$, and let $P_{\mathrm{inc}}(a)$ denote the post-index probability of incident dementia as a function of the running variable $a$ (date of birth). By the continuity assumption of the regression discontinuity design, $P_{\mathrm{prev}}$ is smooth through the cutoff $a_{0}$ and thus equal on both sides. In other words, the eligibility rule can change treatment at $a_{0}$, but it should not create a discontinuous change in the proportion of people who already had dementia before $t_{0}$.

The observed absolute risk difference across the threshold is:

$$\tau_{\mathrm{abs}}=[P_{\mathrm{prev}}+P_{\mathrm{inc}}(a_{0}^{+})]-[P_{\mathrm{prev}}+P_{\mathrm{inc}}(a_{0}^{-})]=P_{\mathrm{inc}}(a_{0}^{+})-P_{\mathrm{inc}}(a_{0}^{-})$$

The cancellation in this simplified expression is conditional. Smooth baseline prevalence is not sufficient on its own, because residual prevalent dementia contributes to the observed outcome only when it is subsequently recorded in hospital data. The cancellation additionally requires eligibility not to change discontinuously the subsequent survival, hospital contact, or probability of post-index hospital coding among residual prevalent cases. Under that assumption, incompletely excluded prevalent dementia can increase the overall level of observed dementia but should not by itself generate an RD discontinuity.

However, the observed relative risk is:

$$RR_{\mathrm{obs}}=\frac{P_{\mathrm{prev}}+P_{\mathrm{inc}}(a_{0}^{+})}{P_{\mathrm{prev}}+P_{\mathrm{inc}}(a_{0}^{-})}$$

which is attenuated toward the null compared with the incident-only relative risk:

$$RR_{\mathrm{inc}}=\frac{P_{\mathrm{inc}}(a_{0}^{+})}{P_{\mathrm{inc}}(a_{0}^{-})}$$

As $P_{\mathrm{prev}}$ increases relative to $P_{\mathrm{inc}}(a)$, $RR_{\mathrm{obs}}$ tends toward 1 even if $RR_{\mathrm{inc}}$ is substantially different from 1. This is directly analogous to non-differential outcome contamination adding the same background component to both sides of a ratio.

A separate issue is the clinically interesting but partly unobservable estimand: the effect of eligibility on incident dementia among people who were truly dementia-free at $t_{0}$. To make this distinction explicit, let $p$ be the probability of residual prevalent dementia at the threshold and let $q_{-}$ and $q_{+}$ be the post-index risks of incident dementia among people without prevalent dementia immediately below and above the threshold, respectively. The observed risk on each side can then be written as

$P_{obs,-}=p+(1-p)q_{-}$ and $P_{obs,+}=p+(1-p)q_{+}$

Therefore the observed absolute RD estimand is:

$$\tau_{\mathrm{obs}}=P_{obs,+}-P_{obs,-}=(1-p)(q_{+}-q_{-})$$

whereas the incident-only absolute estimand among those without prevalent dementia is:

$$\tau_{\mathrm{inc}}=q_{+}-q_{-}$$

Thus,

$\tau_{\mathrm{obs}}=(1-p)\tau_{\mathrm{inc}}$,

and the bias relative to the incident-only absolute estimand is

$\tau_{\mathrm{obs}}-\tau_{\mathrm{inc}}=-p\tau_{\mathrm{inc}}$.

This shows where bias can enter for the incident-only estimand. Under the additional ascertainment assumption stated above, this is not a discontinuity bias caused by prevalent dementia itself; rather, the observed full-population absolute effect is a slightly diluted version of the effect among the baseline dementia-free population. The dilution factor is 1 − p, so the maximum practical importance of this issue is governed by the amount of residual prevalent dementia.

The corresponding expression for the relative risk among all observed participants is:

$$RR_{\mathrm{obs}}=\frac{p+(1-p)q_{+}}{p+(1-p)q_{-}}$$

and, since $RR_{\mathrm{inc}}=q_{+}/q_{-}$, this can also be expressed as:

$$RR_{\mathrm{obs}}-1=\frac{(1-p)q_{-}}{p+(1-p)q_{-}}(RR_{\mathrm{inc}}-1)$$

The multiplier in the expression above is less than 1, so the observed relative risk is pulled toward the null. The attenuation is stronger when residual prevalent dementia is large relative to the incident dementia risk, and weaker when residual prevalent dementia is rare or when the cumulative incident dementia risk is high.

As a simple numerical example, suppose residual prevalent dementia at the threshold were $p=0.03$, the incident dementia risk among those without prevalent dementia were $q_{-}=0.15$ on the ineligible side, and eligibility reduced this to $q_{+}=0.12$ on the eligible side. The incident-only absolute effect would be:

$$\tau_{\mathrm{inc}}=0.12-0.15=-0.03$$

or a 3.0 percentage-point reduction. The observed full-population absolute effect would be:

$$\tau_{\mathrm{obs}}=(1-0.03)(-0.03)=-0.0291$$

or a 2.91 percentage-point reduction. The bias relative to the incident-only absolute estimand would therefore be $0.0009$, or 0.09 percentage points. On the relative scale, the incident-only relative risk would be:

$$RR_{\mathrm{inc}}=0.12/0.15=0.80$$

whereas the observed relative risk would be:

$$RR_{\mathrm{obs}}=\frac{0.03+0.97\times0.12}{0.03+0.97\times0.15}=0.83$$

Thus, smooth residual prevalent dementia is unlikely by itself to create a false RD effect. If eligibility does not discontinuously affect post-index survival, hospital contact, or ascertainment among residual prevalent cases, it does not bias the combined observed absolute estimand. Under the same assumption, it attenuates the absolute effect for the incident-only estimand among people without prevalent dementia at baseline by the factor 1 − p. Relative risk estimates are also attenuated toward 1, but the size of that attenuation is determined by the size of residual prevalent dementia relative to the post-index incident dementia risk.
