## Supplementary Figures for "Eligibility for shingles vaccination and hospital-coded dementia in England and Wales: a regression discontinuity analysis in England"

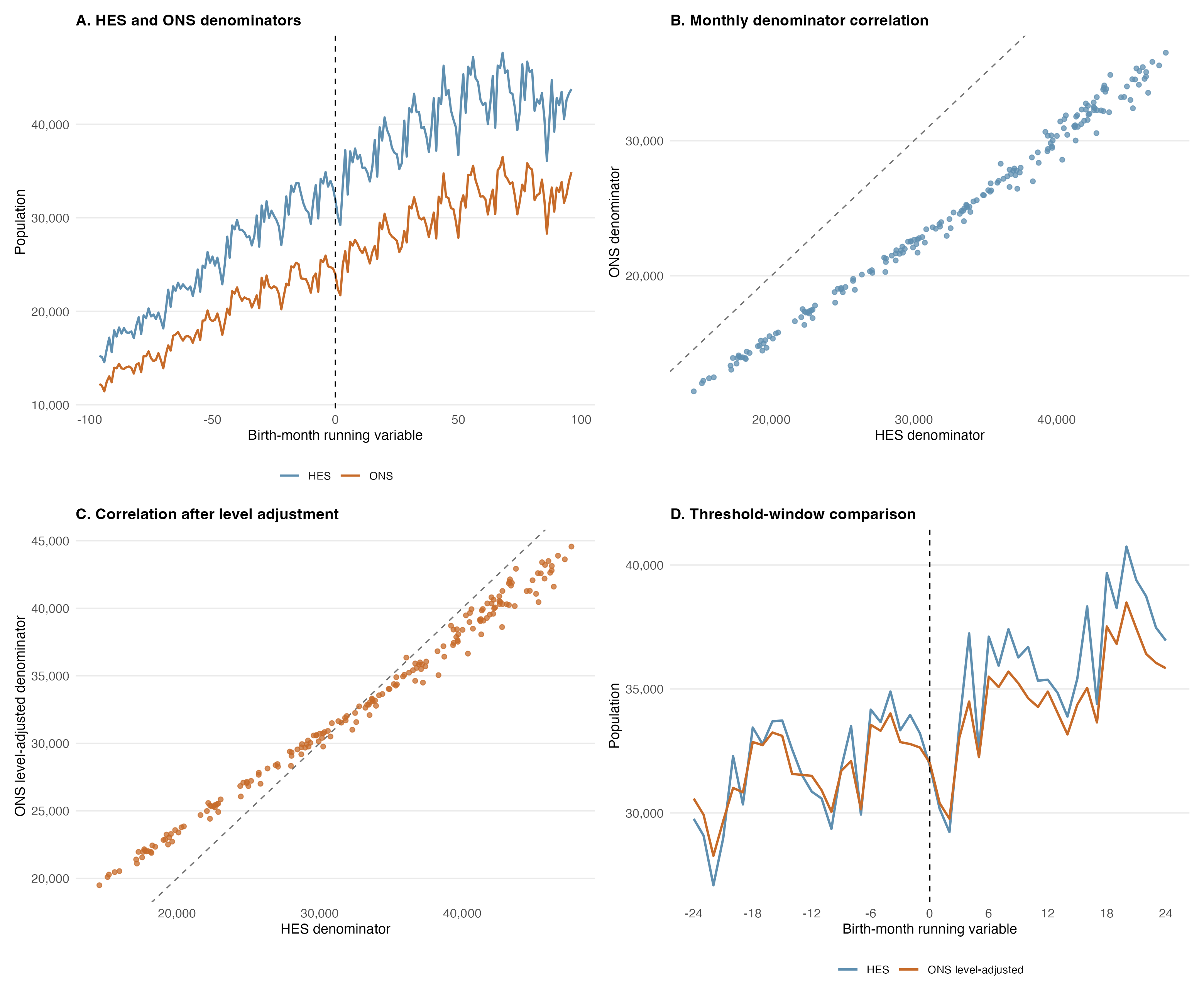

**Supplementary Figure 1**: Comparison of HES and ONS population denominators across the birth-month running variable. Panel A shows monthly denominator counts for the full population using HES and ONS series. Panel B shows the correlation between monthly HES and ONS counts, with each dot representing a month bin, arranged by total population count. Panel C shows the mean-shift corrected ONS series against HES counts. Panel D shows the corrected ONS series and HES series within 24 months of the September 1933 threshold, showing no visible discontinuity.

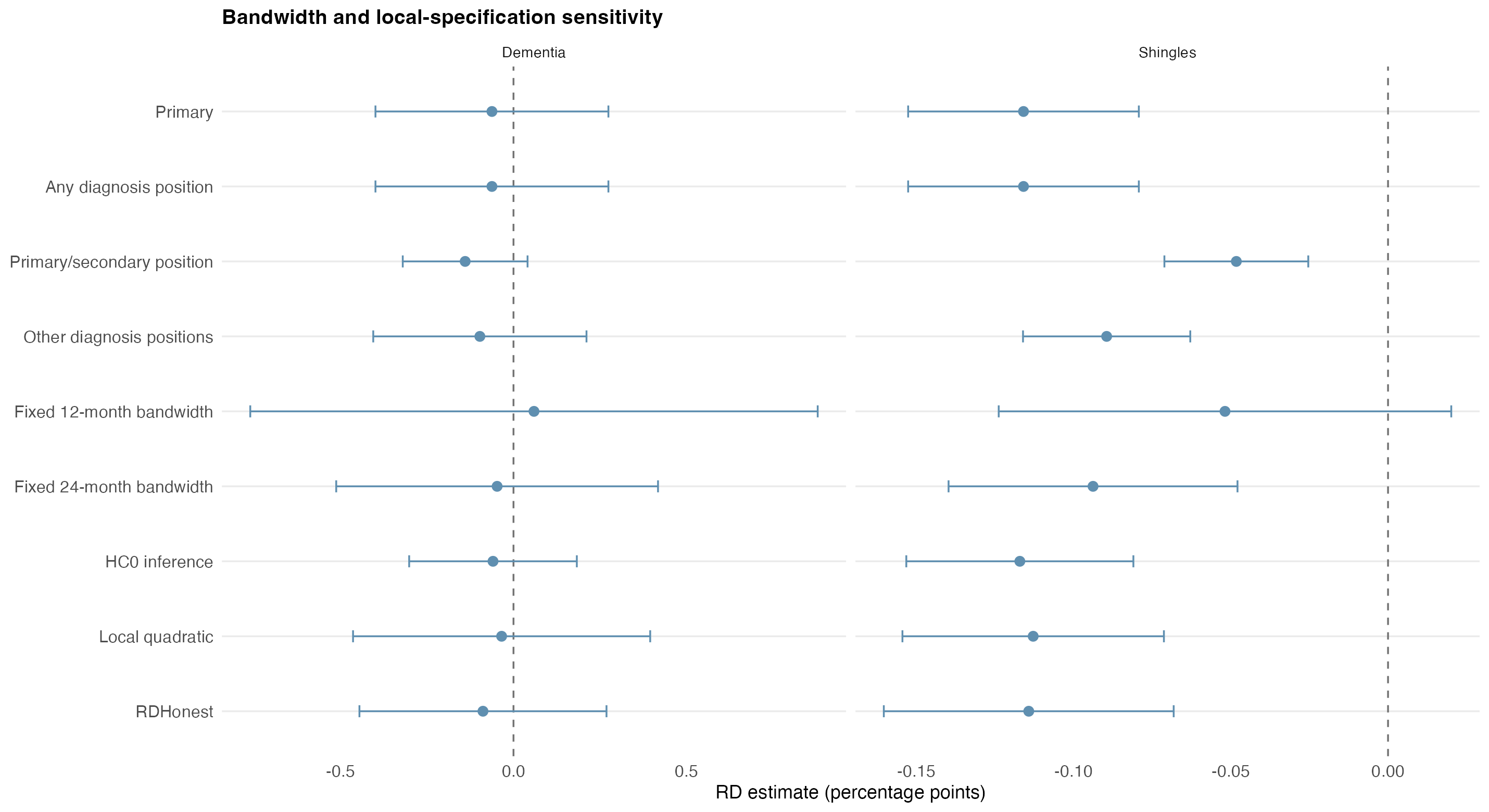

**Supplementary Figure 2:** Sensitivity analyses for the primary shingles and dementia outcomes. Estimates are absolute risk differences with 95% confidence intervals under alternative diagnosis-position definitions, fixed 12- and 24-month bandwidths, HC0 inference, local-quadratic regression, and RDHonest.

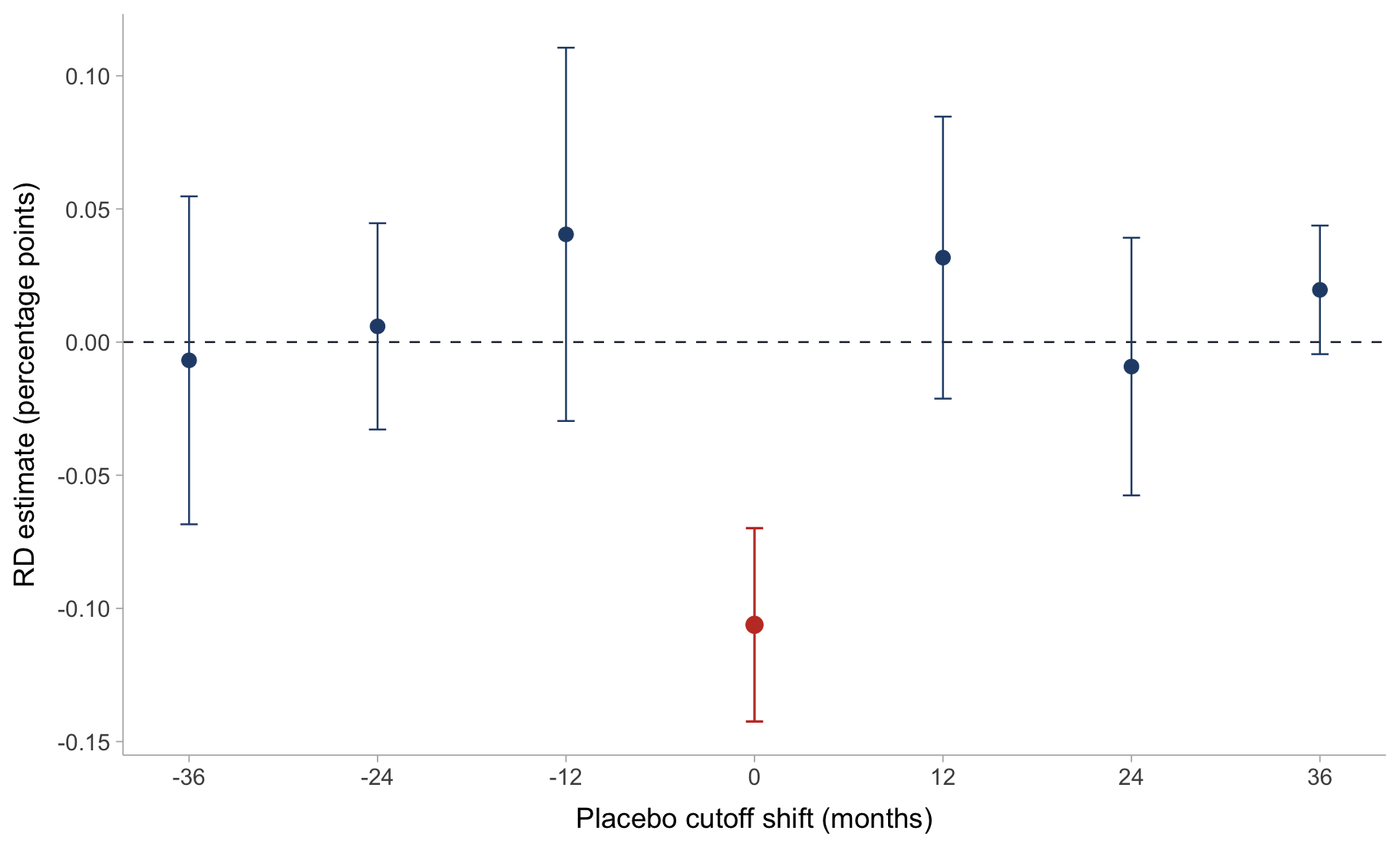

**Supplementary Figure 3:** Annual shingles placebo-cutoff analyses. RD estimates are shown for placebo thresholds shifted by ±12, ±24, and ±36 months around the true September 1933 eligibility cutoff. The true cutoff is shown at shift 0 and highlighted in red.

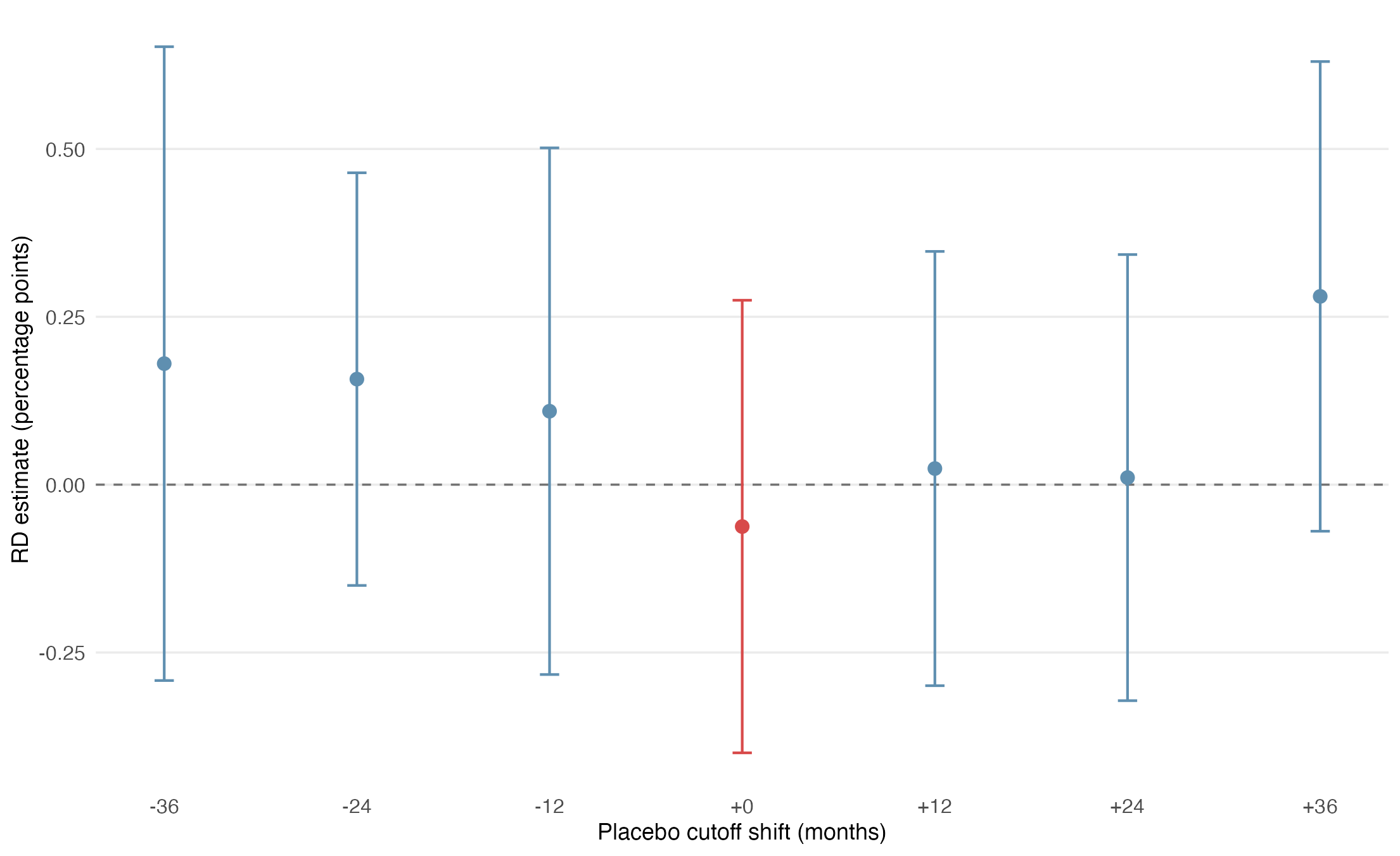

**Supplementary Figure 4:** Annual dementia placebo-cutoff analyses. RD estimates are shown for placebo thresholds shifted by ±12, ±24, and ±36 months around the true September 1933 eligibility cutoff. The true cutoff is shown at shift 0 and highlighted in red. All placebo-cutoff estimates were compatible with the null. The +36-month estimate was the largest positive estimate, but its 95% confidence interval included zero.

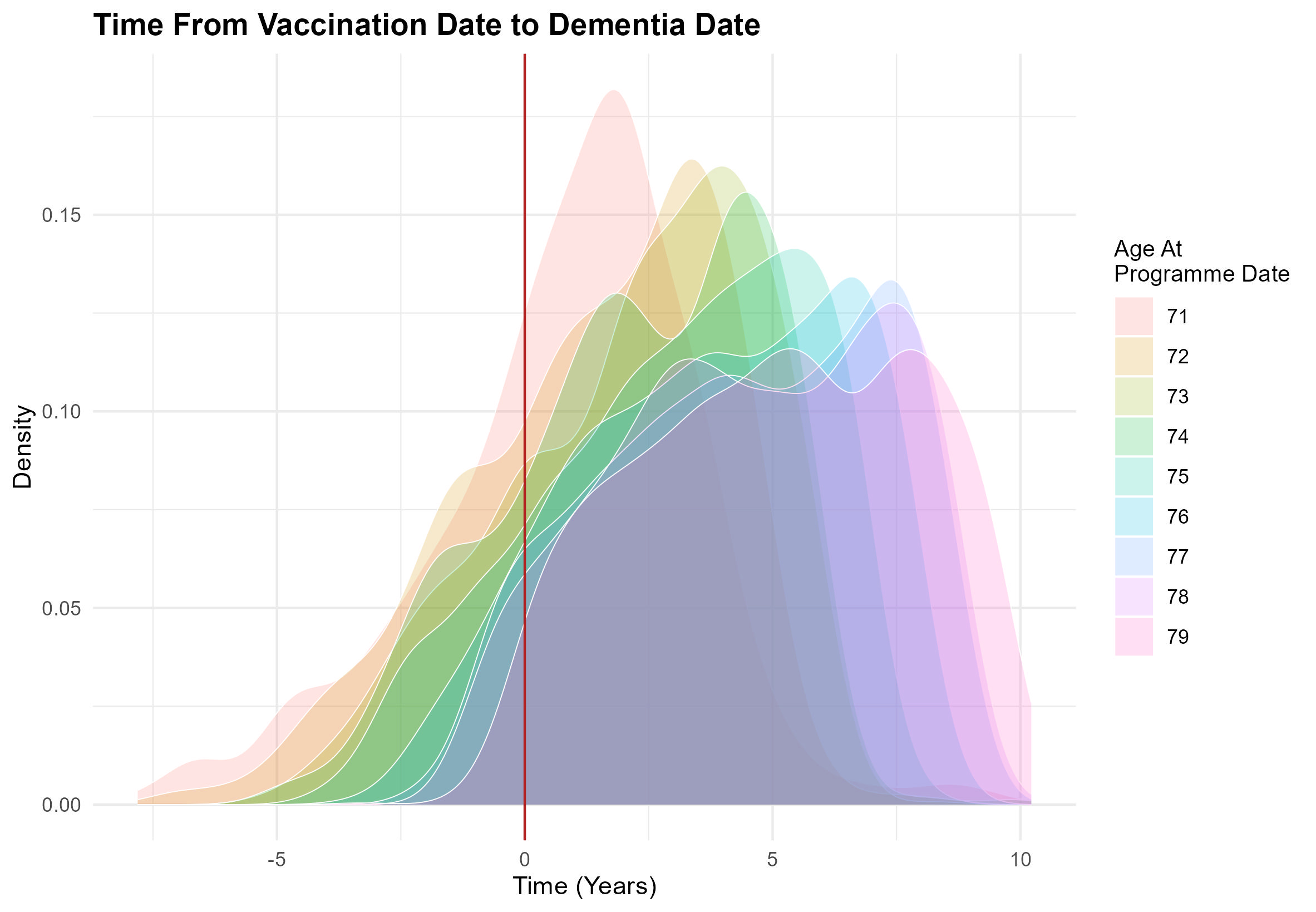

**Supplementary Figure 5: Timing** of shingles vaccination relative to dementia diagnosis in SAIL. Density plots show the interval from vaccination date to dementia date, stratified by age at programme date. Values below zero indicate dementia dates preceding vaccination; the vertical red line marks vaccination date.

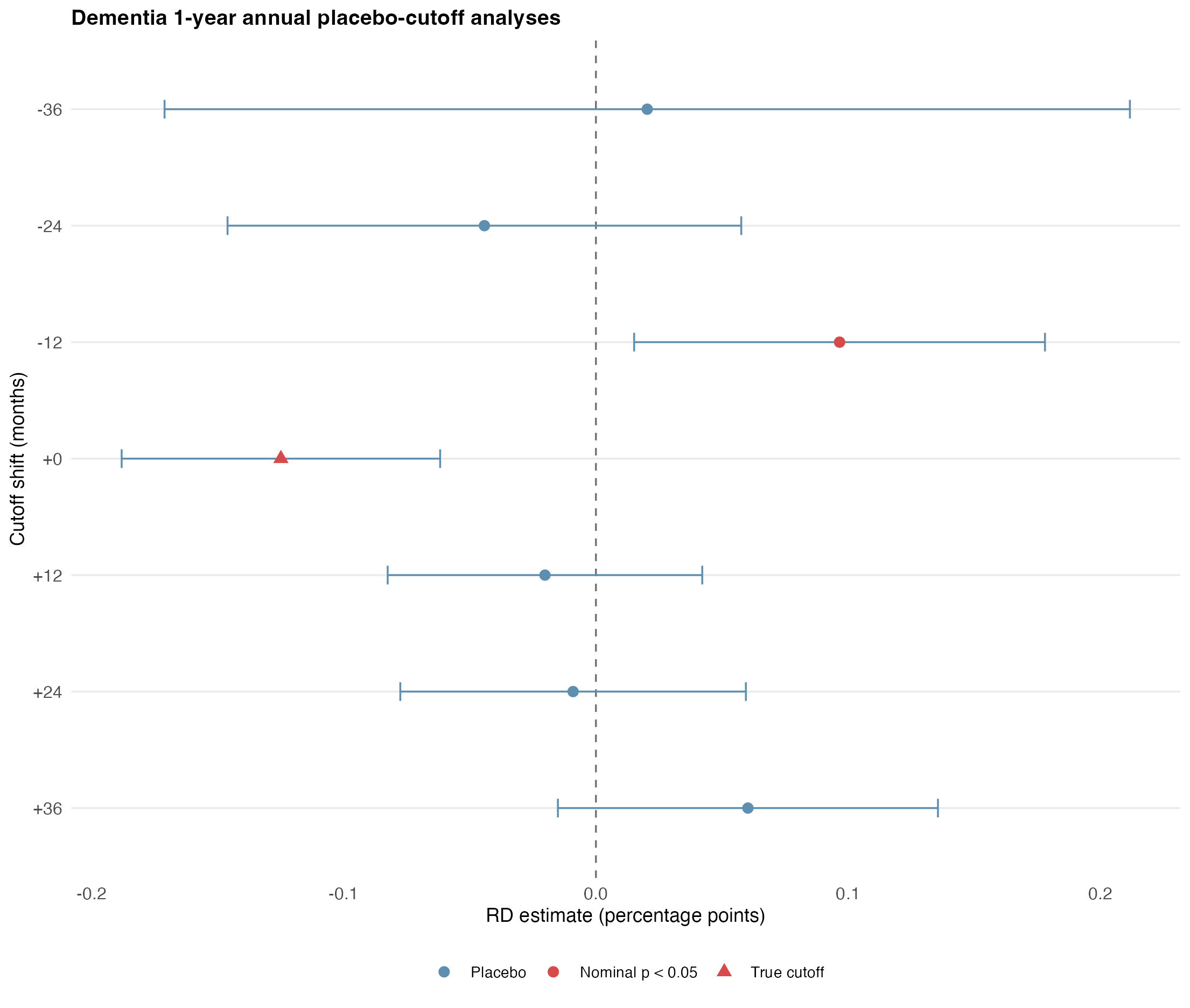

**Supplementary Figure 6:** Annual placebo-cutoff analyses for the 1-year dementia endpoint. RD estimates are shown for annual placebo thresholds shifted by ±12, ±24, and ±36 months around the true September 1933 cutoff, using HES denominators, no grace period, and 1 year of follow-up. The true cutoff estimate is highlighted with a triangle; nominal placebo estimates are highlighted in red. The true 1-year estimate is nominally protective, but a nominal signal is also observed at the -12-month placebo cutoff, arguing against a uniquely cutoff-specific early effect.

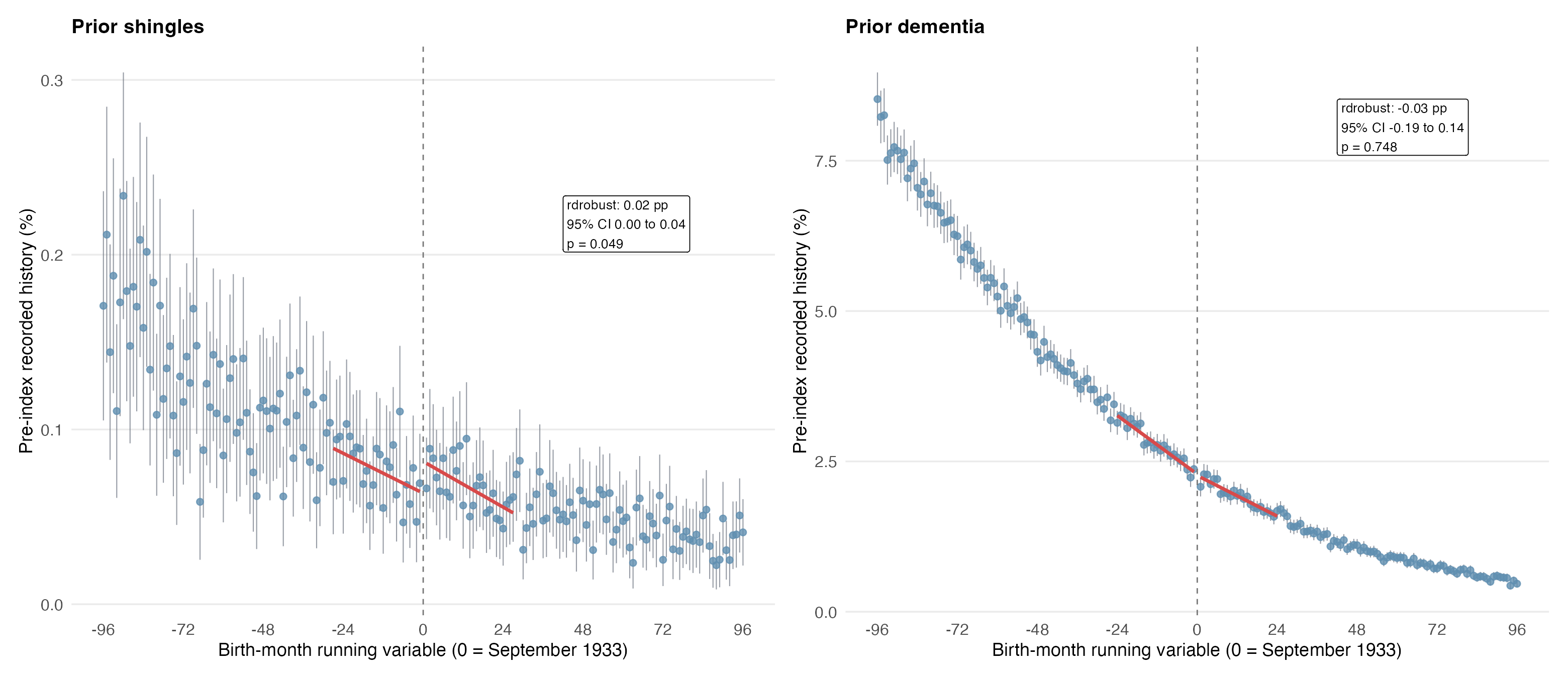

**Supplementary Figure 7:** Pre-index balance checks for the primary outcomes, reflecting outcomes that occurred prior to the RD index date. This figure shows the raw pre-index RD plots for prior shingles and prior dementia.

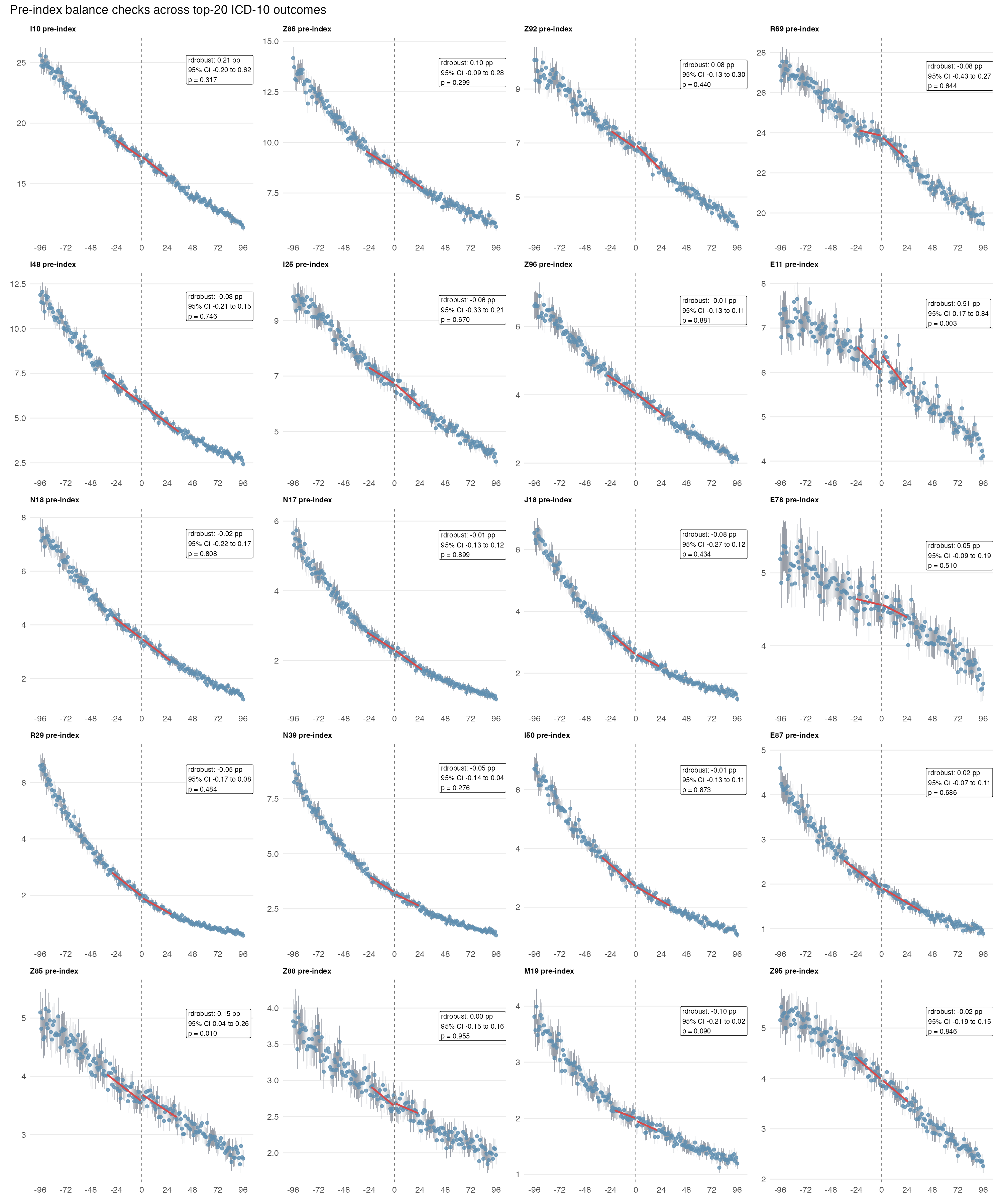

**Supplementary Figure 8:** Pre-index balance checks across the top-20 ICD-10 outcomes. Type 2 diabetes mellitus (E11) shows the clearest raw pre-index imbalance, with additional weaker signals in a small number of other codes. Numerical RD estimates are summarised in Supplementary Table 3.

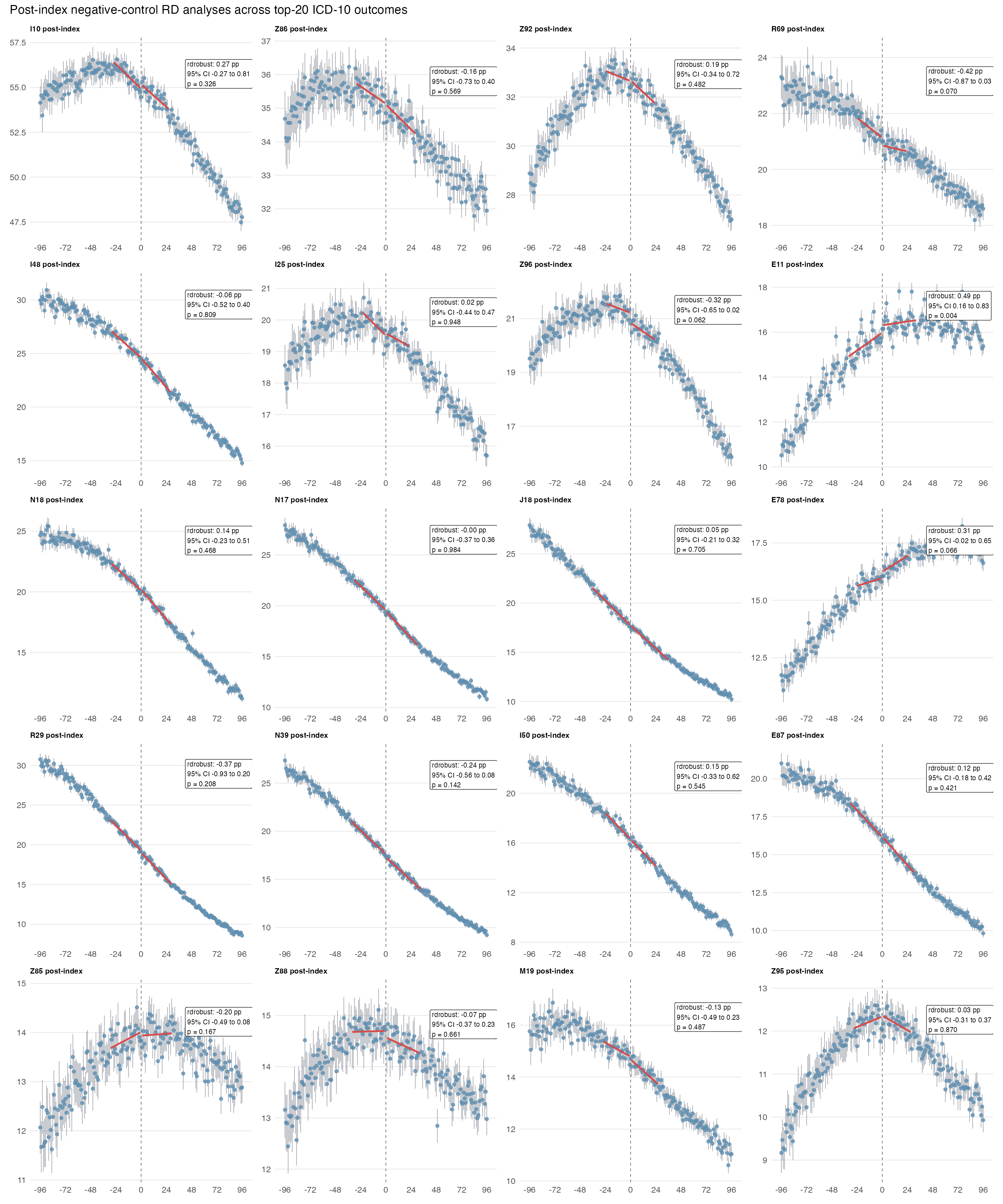

**Supplementary Figure 9:** Post-index negative-control RD analyses across the top-20 ICD-10 outcomes. This figure shows the raw post-index RD plots. Numerical RD estimates are summarised in Supplementary Table 4.

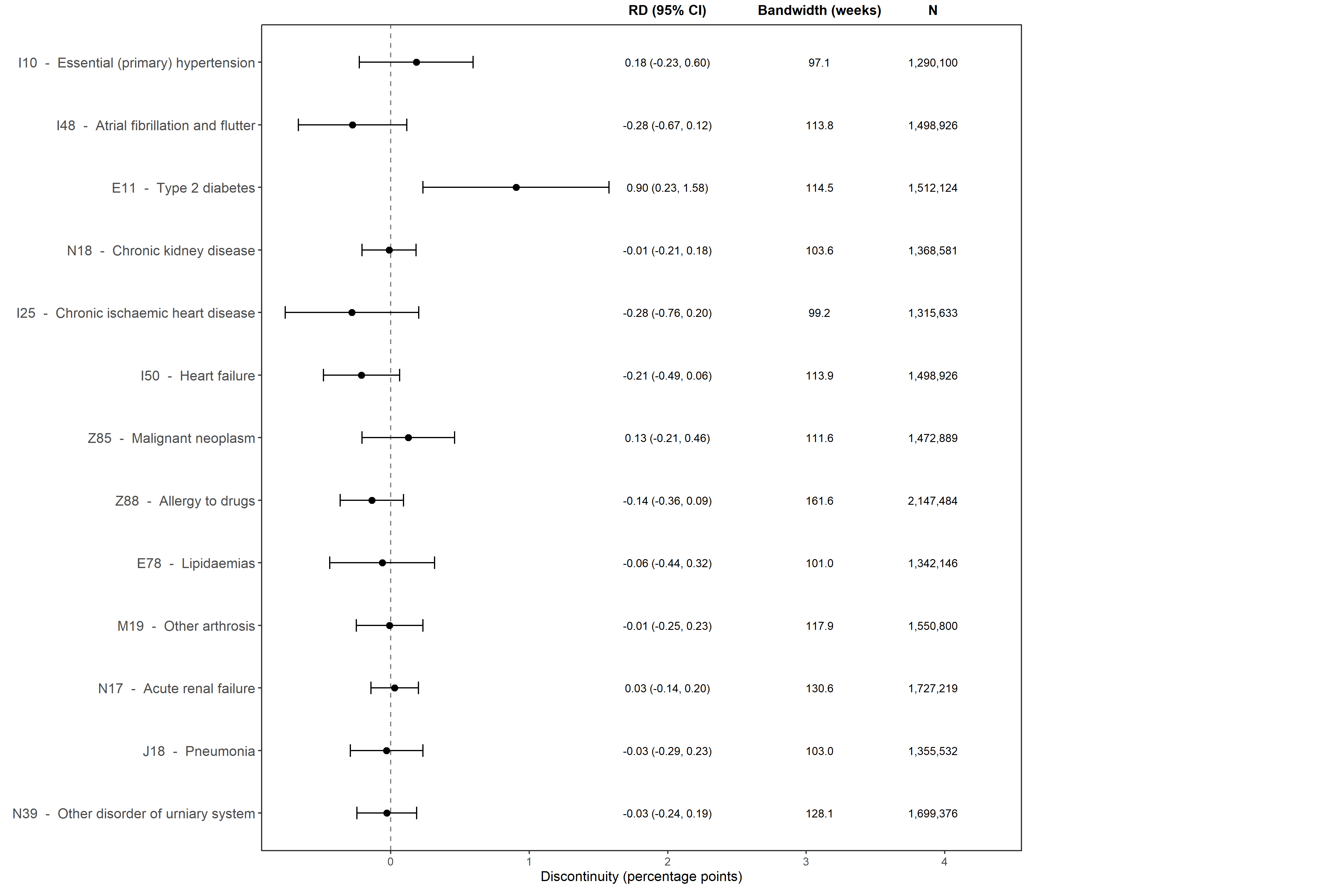

**Supplementary Figure 10:** Pre-index balance checks across common ICD-10 outcomes using approximately 10 years of pre-programme HES history available in the SAHSU HES extract (1 September 2003 - 31 August 2013).

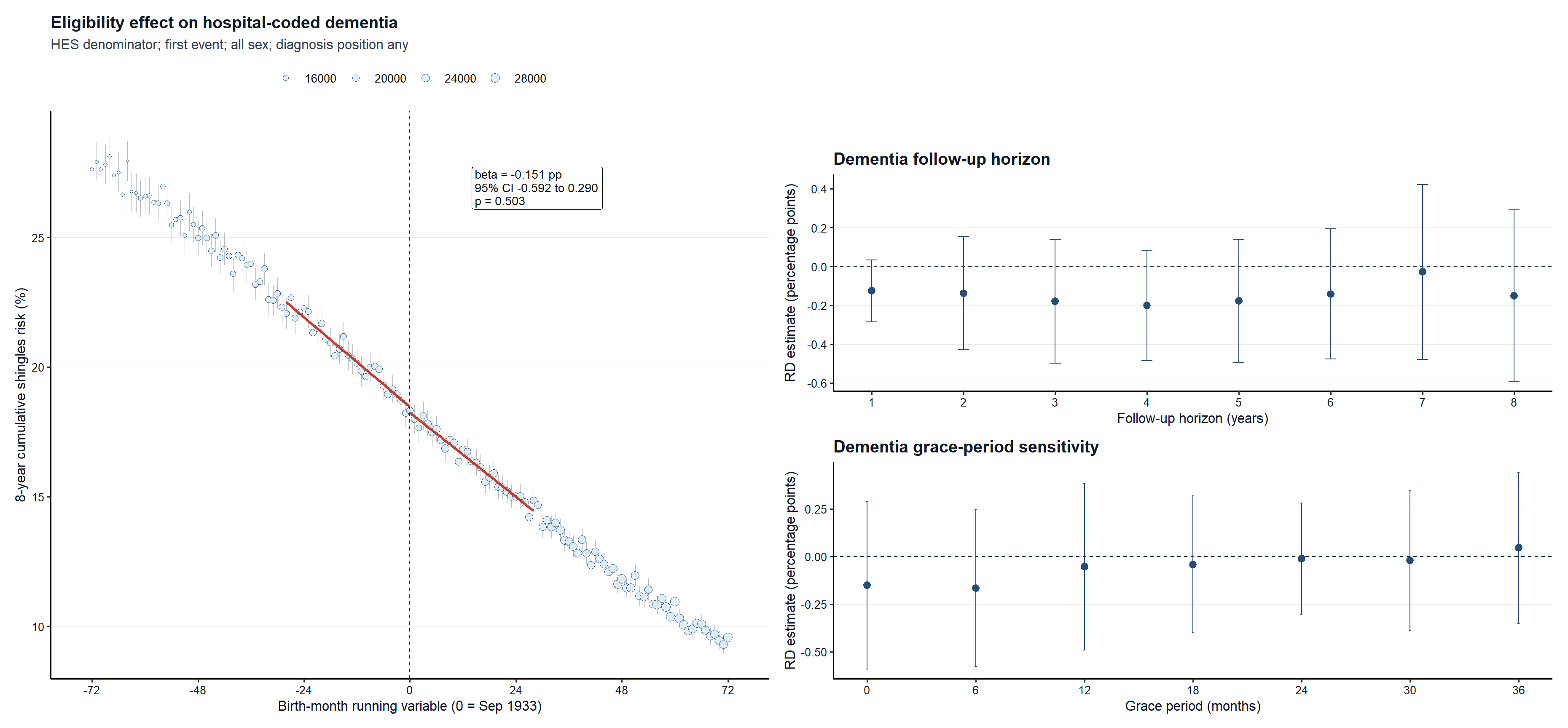

**Supplementary Figure 11:** Effect of eligibility for the shingles vaccination on incidence of dementia following exclusion of prevalent dementia using approximately 10 years of pre-programme HES history (1 September 2003 - 31 August 2013) and a month-of-birth running variable. Panel A shows month-of-birth mean outcome risks with 95% binomial confidence intervals and local-linear fits estimated separately on each side of the eligibility threshold. The outcome is first hospital-coded dementia diagnosis within 8 years among individuals without a hospital-coded dementia diagnosis during the 10 years prior to programme implementation. Panel B shows the effect of increasing follow-up, while Panel C shows the effect of introducing a grace period after the index date before outcome counting begins.

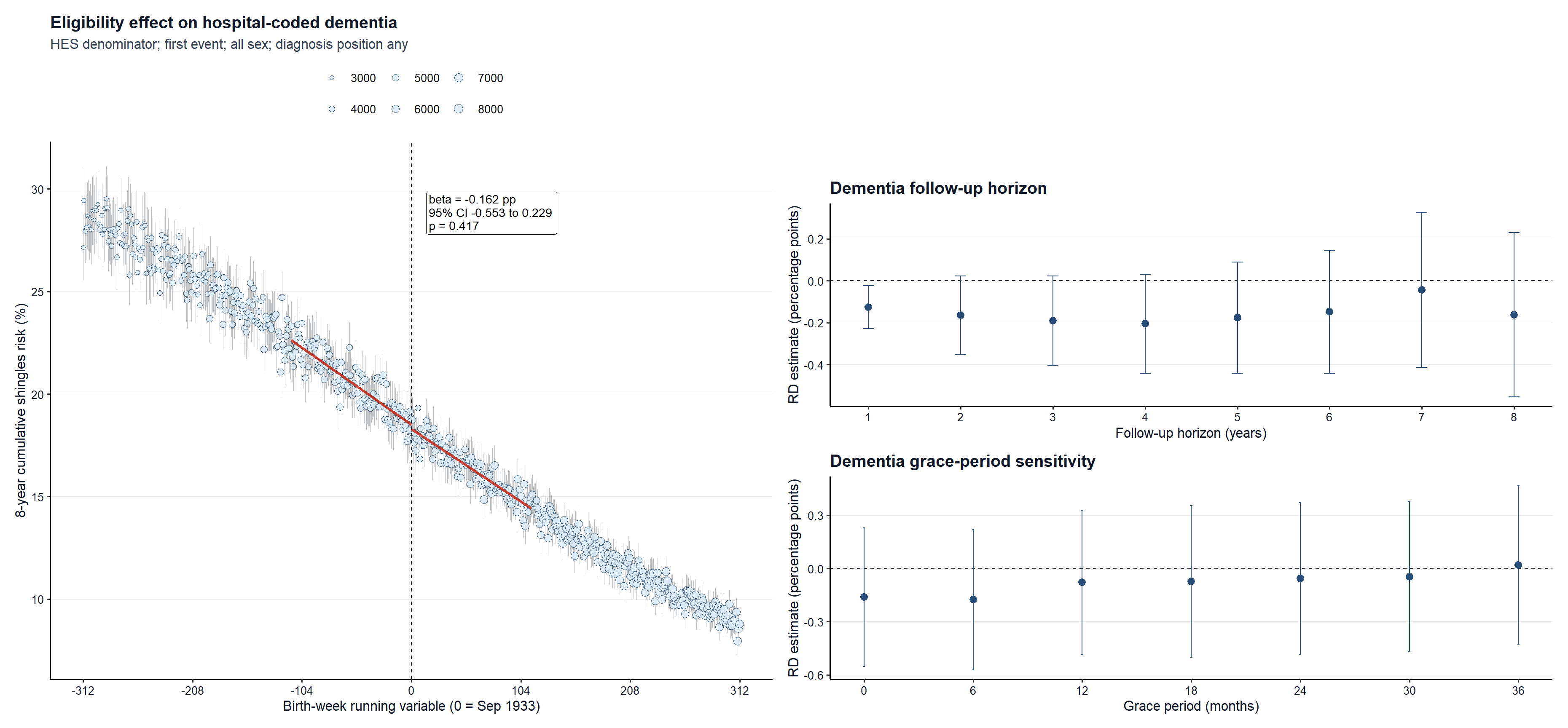

**Supplementary Figure 12:** Effect of eligibility for the shingles vaccination on incidence of dementia following exclusion of prevalent dementia using approximately 10 years of pre-programme HES history (1 September 2003 - 31 August 2013) and a week-of-birth running variable. Panel A shows week-of-birth mean outcome risks with 95% binomial confidence intervals and local-linear fits estimated separately on each side of the eligibility threshold. The outcome is first hospital-coded dementia diagnosis within 8 years among individuals without a hospital-coded dementia diagnosis during the 10 years prior to programme implementation. Panel B shows the effect of increasing follow-up, while Panel C shows the effect of introducing a grace period after the index date before outcome counting begins.

**Supplementary Figure 13:** SAIL all-sex ITT dementia estimates excluding prevalent dementia. Panels show the same all-source versus hospital-only specification matrix as **Figure 6A**, with prevalent dementia excluded. Rows vary SAIL quality control and columns vary the HES-like healthcare-activity restriction; estimates are shown on absolute risk-difference and relative risk-ratio scales.

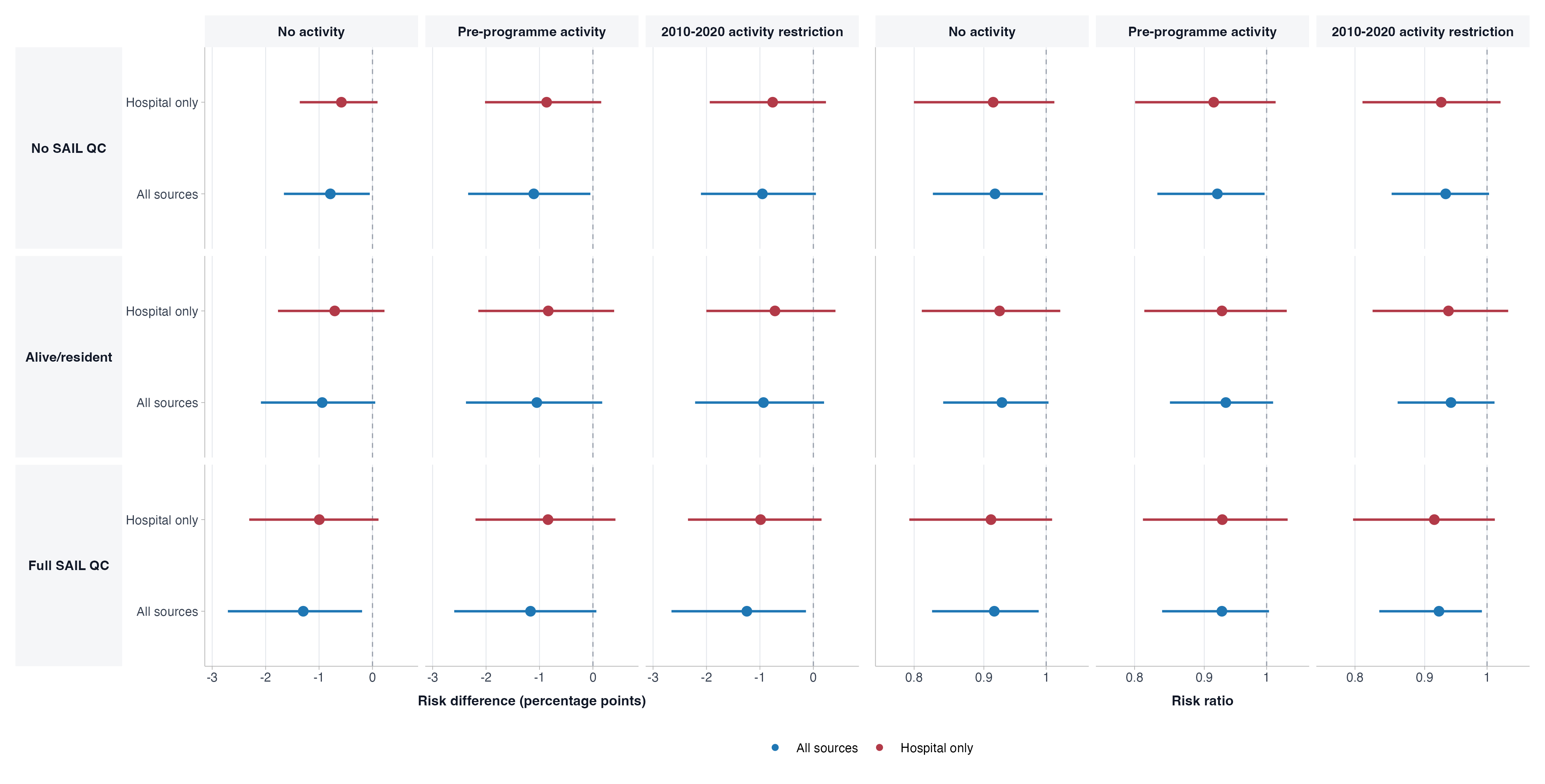

**Supplementary Figure 14:** Sex-specific SAIL ITT dementia estimates including prevalent dementia. Female and male estimates are shown across the same SAIL quality-control, activity-restriction, and ascertainment-source matrix as Figure 6A, with prevalent dementia included.

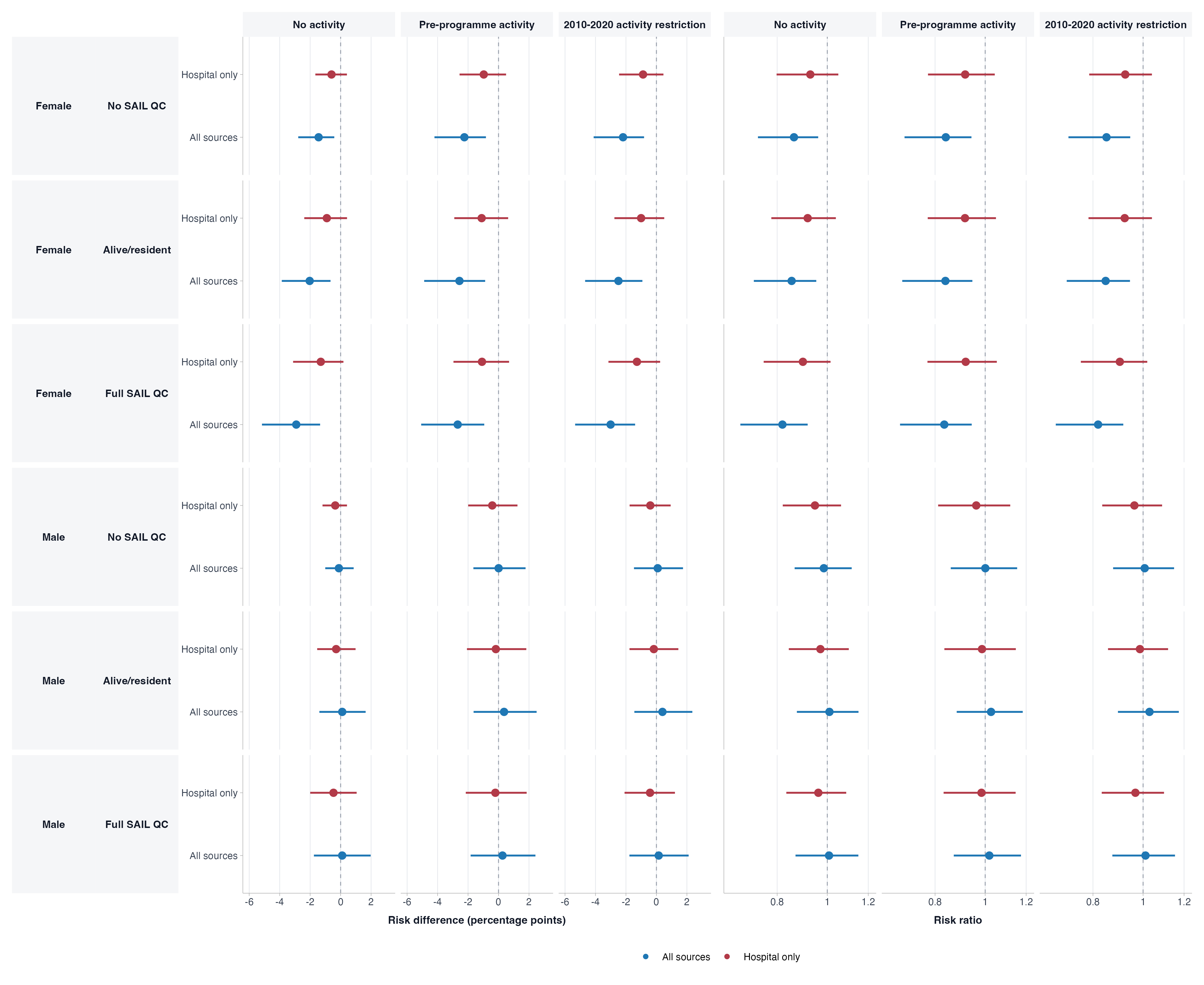

**Supplementary Figure 15:** Sex-specific SAIL ITT dementia estimates excluding prevalent dementia. Female and male estimates are shown across the same SAIL quality-control, activity-restriction, and ascertainment-source matrix as Supplementary Figure 13, with prevalent dementia excluded.

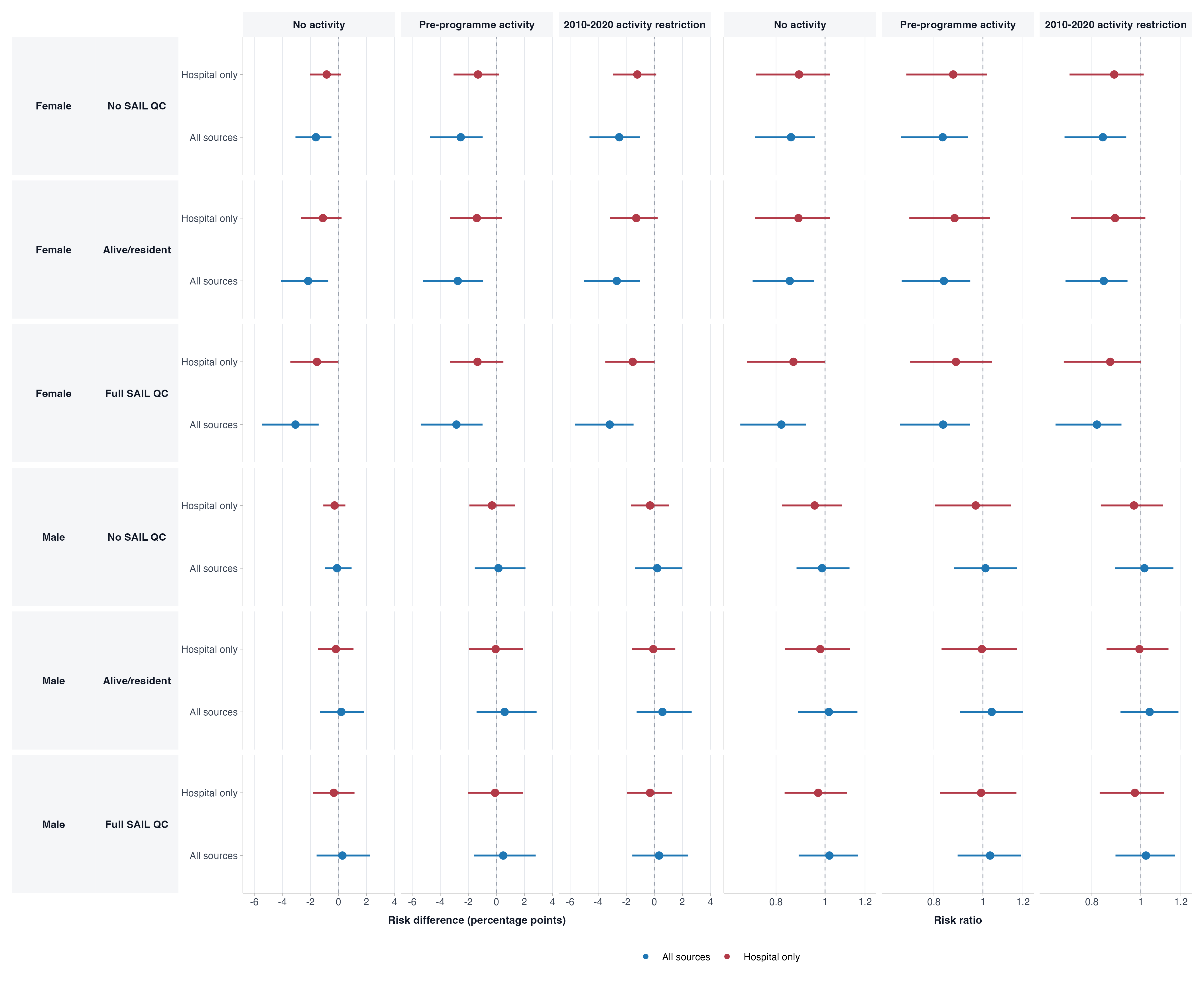
